# Acceptability of app-based contact tracing for COVID-19: Cross-country survey evidence

**DOI:** 10.1101/2020.05.05.20091587

**Authors:** Samuel Altmann, Luke Milsom, Hannah Zillessen, Raffaele Blasone, Frederic Gerdon, Ruben Bach, Frauke Kreuter, Daniele Nosenzo, Séverine Toussaert, Johannes Abeler

## Abstract

**Background:** The COVID-19 pandemic is the greatest public health crisis of the last 100 years. Countries have responded with various levels of lockdown to save lives and stop health systems from being overwhelmed. At the same time, lockdowns entail large socio-economic costs. One exit strategy under consideration is a mobile phone app that traces close contacts of those infected with COVID-19. Recent research has demonstrated the theoretical effectiveness of this solution in different disease settings. However, concerns have been raised about such apps because of the potential privacy implications. This could limit the acceptability of app-based contact tracing among the general population. As the effectiveness of this approach increases strongly with app take-up, it is crucial to understand public support for this intervention.

**Objectives:** The objective of this study is to investigate user acceptability of a contact-tracing app in five countries hit by the pandemic.

**Methods:** We conducted a multi-country, large-scale (N = 5995) study to measure public support for digital contact tracing of COVID-19 infections. We ran anonymous online surveys in France, Germany, Italy, the UK and the US. We measured intentions to use a contact-tracing app across different installation regimes (voluntary installation vs. automatic installation by mobile phone providers), and studied how these intentions vary across individuals and countries.

**Results:** We found strong support for the app under both regimes, in all countries, across all sub-groups of the population, and irrespective of regional-level COVID-19 mortality rates. We investigated the main factors that may hinder or facilitate take-up and found that concerns about cyber security and privacy, together with lack of trust in government, are the main barriers to adoption.

**Conclusions:** Epidemiological evidence shows that app-based contact-tracing can suppress the spread of COVID-19 if a high enough proportion of the population uses the app and that it can still reduce the number of infections if take-up is moderate. Our findings show that the willingness to install the app is very high. The available evidence suggests that app-based contact tracing may be a viable approach to control the diffusion of COVID-19.

## Introduction

The COVID-19 pandemic is the greatest public health threat of the last 100 years. In the absence of effective treatment or vaccination (as of April 2020), the public health response has so far relied on non-pharmaceutical measures to limit the spread of the epidemic, such as physical distancing, case isolation, and manual contact tracing[1]. These measures have not been sufficient to stop the epidemic. Many countries have therefore resorted to partial or full “lockdown” measures to control the epidemic, severely limiting social and economic interactions among their citizens. Although lockdowns may help countries to keep the number of infections under control[2], they come at a great social and economic cost[3–6].

COVID-19 is difficult to trace by traditional methods as COVID-19 cases are infectious 1-2 days before experiencing symptoms and contacts on average become infectious 3-4 days after exposure. The window to achieve containment by manual contact tracing is thus extremely short. Ferretti, Wymant and colleagues[7] have proposed *digital (app-based) contact tracing* as an alternative measure to contain the epidemic without the large economic costs of lockdowns. The idea is to use low-energy Bluetooth connections between phones to record the interactions users have with others, particularly those interactions that may pose a higher risk of infection (e.g., spending more than 15 minutes within two metres of another person). If a user is diagnosed with COVID-19, they can use the app to declare the diagnosis, which leads to a notification of all other users who have come in close contact with the infected person, asking them to isolate at home for 14 days or until they have been tested by the public health authority. The main advantage over traditional (manual) forms of contact tracing is that the app allows *instantaneous* notification of contacts, which is a key determinant of the effectiveness of case isolation and contact tracing strategies for COVID-19[7]. Other advantages are that the automatic recording of contacts scales up easily, and avoids the loss of information due to patients’ recall bias and/or imperfect knowledge of the people they have been in contact with.

In recent weeks, several countries have announced plans to develop various types of contact-tracing apps [8,9], and a few countries have already launched one, e.g., Singapore[10]. The success of app-based contact tracing, however, critically depends on people’s willingness to use the app. Hinch and colleagues[11] simulate the epidemic in the UK and show that the app reduces infections at all levels of take-up but that it is only sufficient to stop the epidemic if approximately 60% of the population use it. It is therefore important to gauge the strength of public support for this approach and to understand the factors that may hinder or facilitate take-up. For instance, since the app would need to trace individuals’ interactions with others, privacy concerns may undermine support and adoption.[12] It is also possible that such a technological solution may not work as well for the less digitally literate share of the population, further increasing the unequal impact of the COVID-19 pandemic within and across countries[13]. In this sense, an “opt-out” installation policy, where mobile phone providers or Apple and Google[14] would automatically install the app on phones, could maximize take-up. It is unclear, however, whether the public would be willing to support this more intrusive solution.

In light of the many open questions surrounding the viability of app-based contact tracing, we designed a survey to measure public support for this approach in five countries that are currently hit by the COVID-19 pandemic: France, Germany, Italy, the UK and the US. The specific objectives of our study are to (i) assess the overall acceptability among the public of app-based contact tracing under different installation policies (e.g., voluntary installation or automatic installation by the government); (ii) uncover country-level and individual-level variation in support for the app; and (iii) understand the main mechanisms that may facilitate or impede app usage across various subgroups of countries and individuals.

## Methods

### Survey design

We conducted large online surveys in five countries (France, Germany, Italy, UK, and the US) to measure acceptability of app-based contact tracing for COVID-19. A complete description of the survey can be found in the Multimedia Appendix; here we provide an overview. At the beginning, after collecting respondents’ informed consent, we described the app, explaining how it would function as well as its purpose. Respondents had to pass a comprehension check to proceed further. We then asked respondents how likely they would be to install the app on their phone, if it became available to download voluntarily (“opt-in” installation policy). Respondents were then asked about their main reasons for and against installing the app as well as their compliance with self-isolation requests. Next, we assessed to what degree respondents would be open to an “opt-out” policy, where mobile phone providers would automatically install the app on all phones, but users would be able to uninstall the app at any time. We then collected demographic information and concluded the survey with questions about respondents’ attitudes towards the government under different installation regimes.

We kept the survey design as similar as possible across all five countries, with a few exceptions to accommodate country differences with regard to lockdown measures in place at the time of taking the survey. The US survey (deployed last) contained a few additional questions, including robustness checks. See Section A in the Multimedia Appendix for more details.

Ethics approval was obtained from the University of Oxford (reference number ECONCIA20-21-06). No personal information was collected as part of the study.

### Target population, sample size and attrition

The surveys were administered between 20^th^ March and 10^th^ April 2020. We recruited respondents through Lucid, an online panel provider. We targeted a sample size of 1000 respondents in each of the four European countries, and 2000 in the US – with quotas set for the samples to be representative of the overall population in terms of gender, age and region of residence. A total of 10375 individuals started the survey and 10308 consented to participate (a participation rate of 99%). Out of the people who consented to participate, 6166 passed the comprehension check and started the main questionnaire. After removing incomplete responses and duplicates, we have a sample of 6061 complete and unique responses (a completion rate of 59%). Finally, we removed 66 respondents who either did not own a mobile phone or did not disclose their gender, leaving us with a final sample of 5995 respondents. To control for the potential effect of our recruitment method, we repeated the German survey with a probability-based sample in an offline recruited online panel. See Section B in the Multimedia Appendix for further details on recruitment, filtering and attrition, and the final sample.

### Statistical analysis

Our main outcome variables measure respondents’ intention to have the app installed on their phone under the two installation regimes (opt-in vs. opt-out). The outcomes were measured on a 5-point ordinal scale (opt-in: from *Definitely install* to *Definitely won’t install*; opt-out: from *Definitely keep* to *Definitely uninstall*). In our regression analysis, we dichotomize these outcome measures (= 1 if definitely or probably install / keep the app, and = 0 otherwise).

We use multivariate regression analysis (linear probability models; probit and ordered logit in additional analyses presented in the Multimedia Appendix) to examine the relationship between intention to install and a number of covariates: age, gender, country, presence of comorbidities (diabetes, high blood pressure, heart or breathing problems), usage of mobile phone outside the house, frequency of social interactions, ability to work from home during the lockdown, ability to obtain sick pay while working from home, trust in national government, and incidence of COVID-19 deaths in a respondent’s region of residence (see Section C.3 in the Multimedia Appendix for more details). Table B.2 in the Multimedia Appendix presents a summary of these covariates.

## Results

### Result 1

*We find broad support for app-based contact tracing. Support is high in all countries, across all subgroups of the population, and under both installation regimes (opt-in and opt-out)*.

Panel A of Figure 1 shows that, under the voluntary (opt-in) installation regime, 74.8% of respondents across all countries would probably or definitely download the contact-tracing app, if it was available. Panel B shows that 67.7% of respondents would also probably or definitely keep the app installed on their phone under the automatic (opt-out) installation regime. In both regimes, the share of respondents who would not have the app installed on their phone is very small (red portion of the bars in Figure 1).

**Figure 1:**
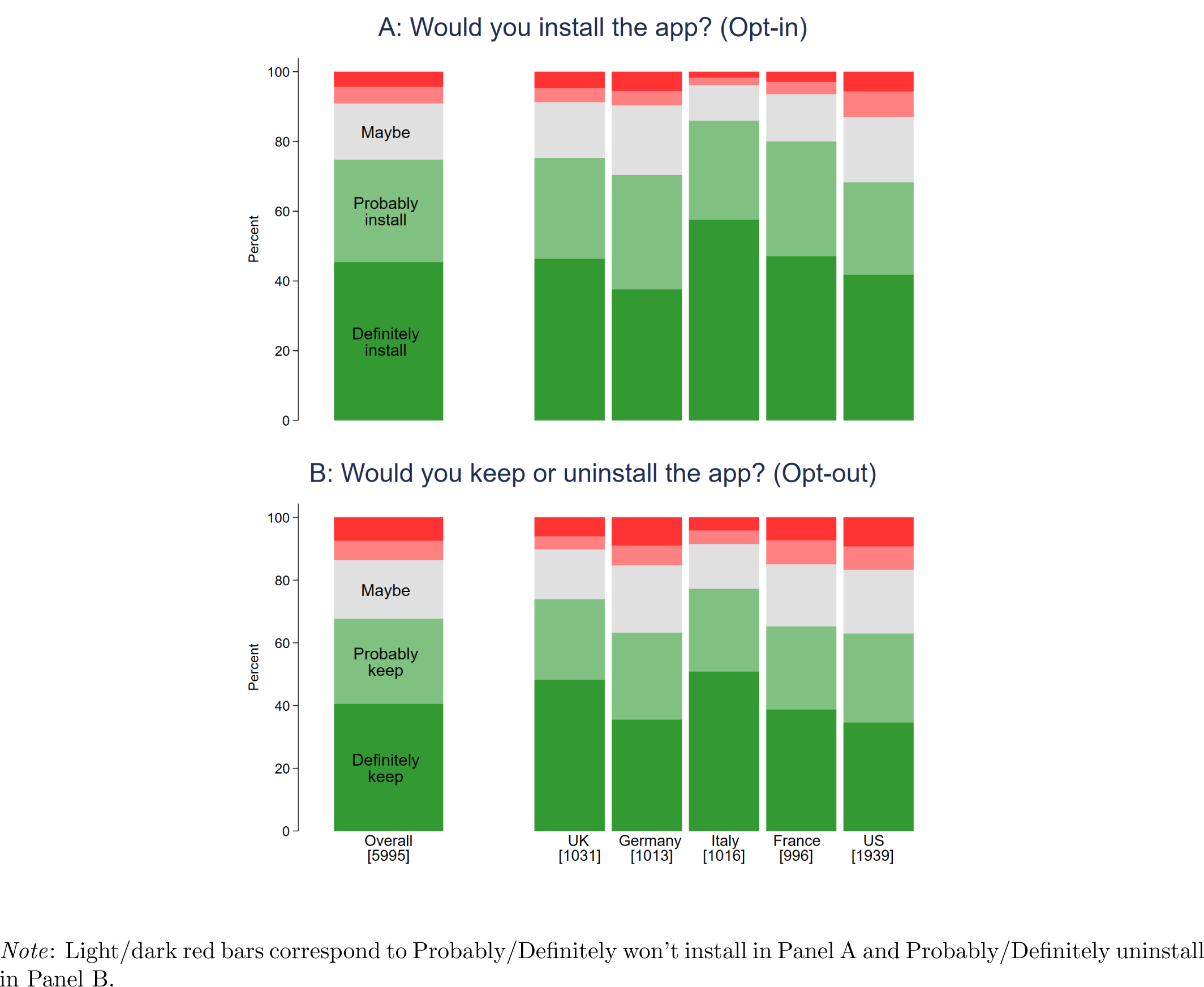
Likelihood of having the app installed, under opt-in and opt-out regimes and by country

**Figure 2:**
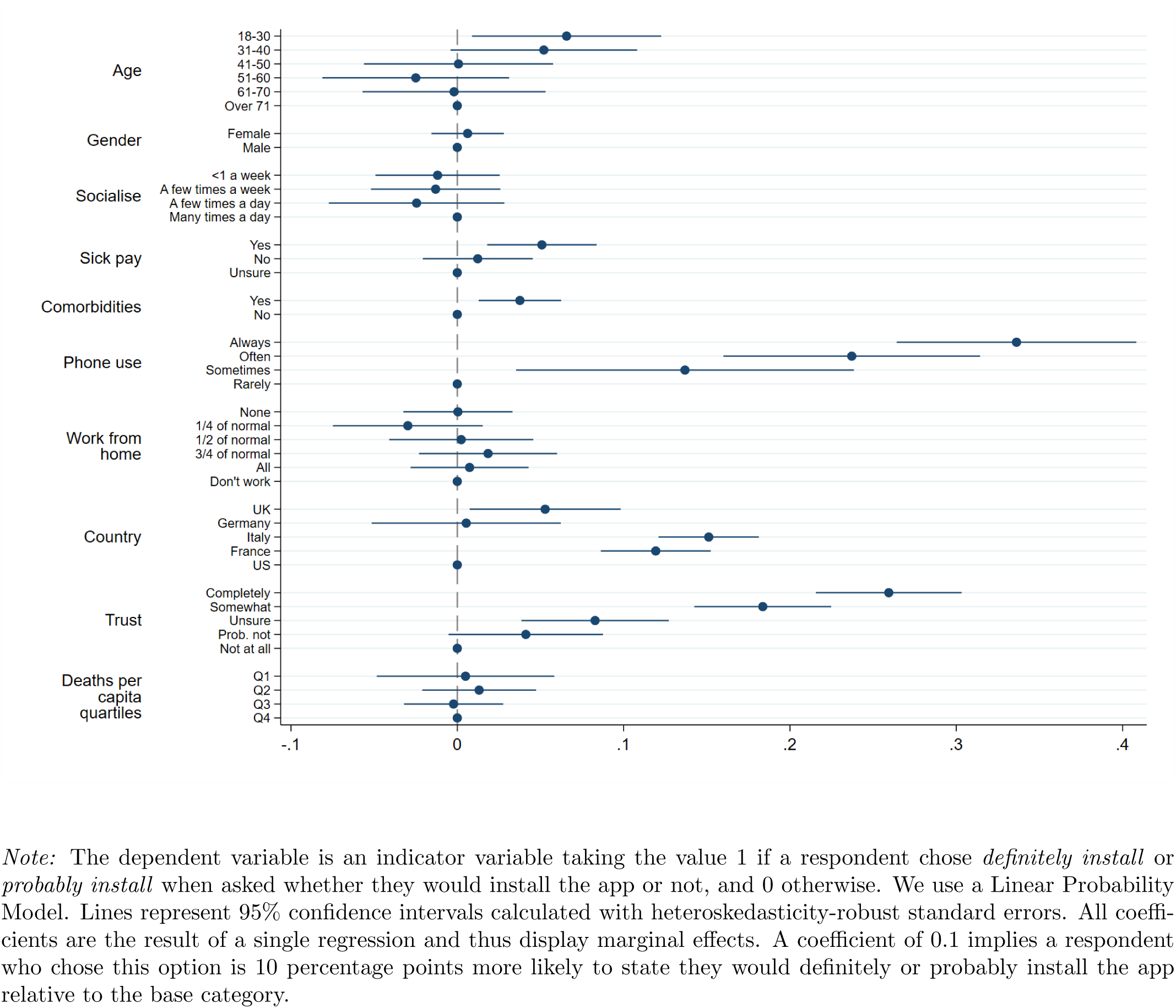
Determinants of stating *definitely install* or *probably install*

Support is high in all five countries where we implemented the survey: in each country, at least 68% of respondents say that they would install or keep the app. Moreover, Figures 9, 10, and 11 in the Multimedia Appendix show that support for the app is generally high across various subgroups of the population (e.g., across men and women, across different age groups, etc.), suggesting widespread acceptability of the app-based contact tracing solution to the COVID-19 pandemic.

### Result 2

*Although support is very high overall, there is systematic variation both within and across countries*.

Despite the broad and widespread acceptability of the app, we find that support varies systematically across countries and individuals. For instance, Figure 1 shows that Germany and the US are relatively less supportive of the app compared to the other countries. This is the case both under the opt-in and opt-out regimes. Among individual characteristics, we find that those who have lower trust in their national government are more hesitant to have the app installed on their phones (Figure 11 in Multimedia Appendix).

We further explore this heterogeneity using multivariate regression analysis, where we examine the relationship between support for the app and a variety of individual- and country-level covariates. Figure 2 shows the impact that these covariates have on the probability of definitely or probably installing the app under the opt-in regime, using a linear probability model (see Section C.1 in the Multimedia Appendix for a similar analysis of the opt-out regime).

The analysis confirms that Germany and the US are significantly less supportive of the app, especially compared to France and Italy. Taking the two most extreme cases, respondents in Italy are 15.1 percentage points (95% CI 12.1 - 18.1) more likely to support the app than respondents in the US. Surprisingly, Figure 2 shows very little correlation between regional-level COVID-19 mortality rates and support for the app.

Among individual-level characteristics, we find that people who carry their phone with them more often are more likely to install the app. Those who always carry their phone with them are 33.6 percentage points (95% CI 26.4 - 40.8) more likely to support the app than those who carry their phone only rarely. App support is also 3.7 percentage points (95% CI 1.3 - 6.2) larger among respondents with one or more comorbidities. Moreover, the probability of installing the app increases with trust in the government. People who completely trust the government are 25.9 percentage points (95% CI 21.6 - 30.3) more likely to install the app than those who do not have any trust in the government.

We find similar results using an ordered logit model, a linear probability model dichotomizing on just *Definitely install*, and when using a probit model (see Multimedia Appendix). Finally, results are also qualitatively similar when considering installation intentions under opt-out rather than opt-in (Figure 8 in Multimedia Appendix). Interestingly, under the opt-out regime, trust in government displays an even stronger correlation with the intention to keep the app installed on one’s phone.

### Result 3

*Concerns about privacy and app security underlie some of the key variation in support for the app*.

We can use the data on respondents’ reasons for or against installing the app to better understand the nature of the observed variation in app support across countries and individuals. A first set of reasons against the app revolved around concerns about government surveillance at the end of the epidemic (mentioned by 42% of respondents) and cyber security (fears that the app could make the phone vulnerable to hackers; 35%). Respondents also reported that usage of the app may increase feelings of anxiety (26%), possibly reflecting aversion to feedback about a possible infection. The most frequent reasons in favor of the app were willingness to protect family and friends (68%), a sense of responsibility towards the community (53%), and a hope that the app may stop the epidemic (55%). Figures 16 and 17 in the Multimedia Appendix show the relationship between the probability of selecting a particular reason and country- and individual-level characteristics.

Several patterns are of interest. First, we find that, compared to other countries, respondents in Germany and the US are more likely to mention concerns about government surveillance as one of the reasons against installing the app. In these countries we also see a larger share of respondents expressing concerns about security of the app, especially compared to Italy and the UK. Thus, concerns about privacy and security seem to be an important impediment to the adoption of the app, particularly in Germany and the US.

Among individual-level characteristics, we find that respondents who have less trust in their national government are also more likely to express concerns about government surveillance. This suggests that privacy concerns play a role in the negative relationship between trust in government and probability of installing the app found in Figure 2. In contrast, we find that frequent usage of mobile phones is related to a stronger perception of the potential benefits of the app: respondents who more often carry their phone with them are more likely to believe that the app would benefit them, by helping them stay healthy and keeping them informed about the risks of infection.

## Discussion

### Principal Findings

In our study, we find high support for app-based contact tracing – irrespective of age, gender, region or even country of residence. Since the effectiveness of app-based contact tracing crucially depends on a sufficient level of take-up, our findings are encouraging for the prospects of this approach. Although support is high in all countries and subgroups of the population, the data reveal that concerns about cyber security and privacy, coupled with trust in government, are important determinants of support. Countries with stronger privacy and security concerns (Germany and the US) are relatively less supportive of app-based contact tracing. Individuals who have less trust in their national government are also less supportive.

### Implications

The lack of trust in government can have far-reaching implications. Our analysis shows that this factor has a negative effect on people’s intention to install a contact-tracing app on their phones. Furthermore, supplementary analysis (see Section C.6 in the Multimedia Appendix) also shows that people with lower trust in government are more in favor of an opt-in installation policy than an opt-out regime where the government asks mobile phone providers to automatically install the app on all phones. An opt-out regime is likely to translate into higher effective installation rates, for instance by reducing the negative effects of procrastination or inattention[15]. However, our data suggests that only governments that enjoy a relatively high level of trust from their citizens may be able to resort to more paternalistic approaches. A policy implication of these findings is that governments should consider delegating the organization of app-based contact tracing to a highly-reputable and transparent public health authority at arm’s length from the government.

Our results also point towards the need to address privacy and cyber security concerns with an app design that respects user personal data as much as possible. Research on the privacy implications of app-based contact tracing, and the potential solutions to these concerns, is currently underway[12,16,17]. Interestingly, however, when we ask our respondents how the data collected by the app should be treated, we find that nearly 60% would consent to making the de-identified data available to research.

### Limitations

Our study has some limitations that we tried to address in different ways. First, respondents recruited online may not be representative of the entire population. In particular, digital literacy and willingness to share data could be higher among such respondents. To ensure that our results do not hinge on our specific sample, we replicated an abridged version of the German survey with a different panel provider that randomly recruits its participants offline. Our results remain almost completely unchanged (see Section B.3 in Multimedia Appendix).

Second, our survey asked hypothetical questions about future behavior. However, high levels of *intended* installations may not directly translate into *actual* installations. Nevertheless, studies often find good correlation between what people declare they would do in surveys and actual behavior[18–22], even in relation to app installations[23–26]. More generally, broad support for the approach is a necessary first stage to adoption, and our findings about heterogeneity in support point towards specific subgroups of the population that may need stronger encouragements to adoption. We show in Section C.4 of the Multimedia Appendix that respondents who would install the app mention far more reasons *for* its adoption than those who would not install it (but a similar number of reasons *against*). We show in Section C.9 that respondents in our replication study who did not answer the comprehension questions correctly were less willing to install. Stressing the various benefits of the app, to oneself and others, and explaining the working and purpose of the app may be a particularly effective strategy to foster adoption.

Third, in our survey, we measured support for the general concept of app-based contact tracing, leaving out specific details regarding the implementation, which were not available to us at the time respondents took the survey. One downside of only surveying about the general idea is that it might be harder for respondents to visualize how such a system could work, which may increase hypothetical bias. However, we find that the details we gave about implementation (e.g., whether the app uses Bluetooth or GPS) seem to have very little impact on support. This suggests that our general measure of support for app-based contact tracing may be portable across different implementation settings.

Finally, our survey respondents were recruited from a specific subset of industrialized Western democracies. Attitudes towards app-based contact tracing may vary across countries with different levels of development and political regimes. It is nevertheless encouraging, in terms of external validity, that we observe a strong similarity in responses across the five countries we sampled, and that analogous findings have been reported in ongoing surveys conducted in Australia and Taiwan[27]. In developing countries and among disadvantaged populations, the more limited access to smartphones raises both efficacy and equity issues; the development of low-cost Bluetooth devices with similar functionalities could improve access to digital contact tracing.

### Conclusions

In conclusion, our study shows strong public support for app-based contact tracing to tackle COVID-19. This is an important result since public support is a necessary condition for the viability of the approach. Further research is needed to gauge the extent to which public support for app-based contact tracing translates into actual app adoption and, more generally, to evaluate its potential for epidemic control.

## Data Availability

All data are available at https://osf.io/7vgq9/ under a CC-BY license.

## Acknowledgements

We acknowledge funding from the Economic and Social Research Council (grant ES/R011710/1), the University of Oxford and Volkswagen Foundation (grant “Consequences of Artificial Intelligence for Urban Societies”).

## Authors’ contributors

Samuel Altmann: figures, study design, data collection, data analysis, writing

Luke Milsom: figures, study design, data collection, data analysis, writing

Hannah Zillessen: study design, data collection, data analysis, writing

Raffaele Blasone: literature search, data collection

Frederic Gerdon: data collection, data analysis

Ruben Bach: data collection, data analysis

Frauke Kreuter: study design, data collection, data analysis

Daniele Nosenzo: study design, data collection, data analysis, writing

Séverine Toussaert: study design, data collection, data analysis, writing

Johannes Abeler: study design, data collection, data analysis, writing

## Abbreviations

COVID-19: coronavirus disease 2019

95% CI: 95% confidence interval

## Multimedia Appendix

Survey details, additional results and survey questionnaire.

## Multimedia Appendix

### A Survey Details

#### A.1 Survey Design

Table 1 gives an overview of the structure of the survey that was administered to respondents. In the following, we describe the various parts in more details. After expressing their informed consent to take part in the study, respondents were given a description of the contact tracing app. We explained that the app would be developed by a national health organization and that, once installed, it would register other users in close proximity via Bluetooth (and, in the case of the US and the UK, potentially location data). Users who were found to have been in close proximity to a confirmed case of COVID-19 for at least 15 minutes would be alerted by the app and asked to quarantine at home for 14 days or until they could be tested for the virus. All other users would see an “all clear” message. We further explained that an early quarantine would prevent individual users from passing on the virus to their loved ones in the early (presymptomatic) stages of their potential infection, and might slow down or even stop the epidemic. We also stressed that the identity of all users would be private throughout the entire process. In order to progress to the main part of the survey, respondents had to correctly answer three comprehension questions about the functioning of the app.

**Table 1:**
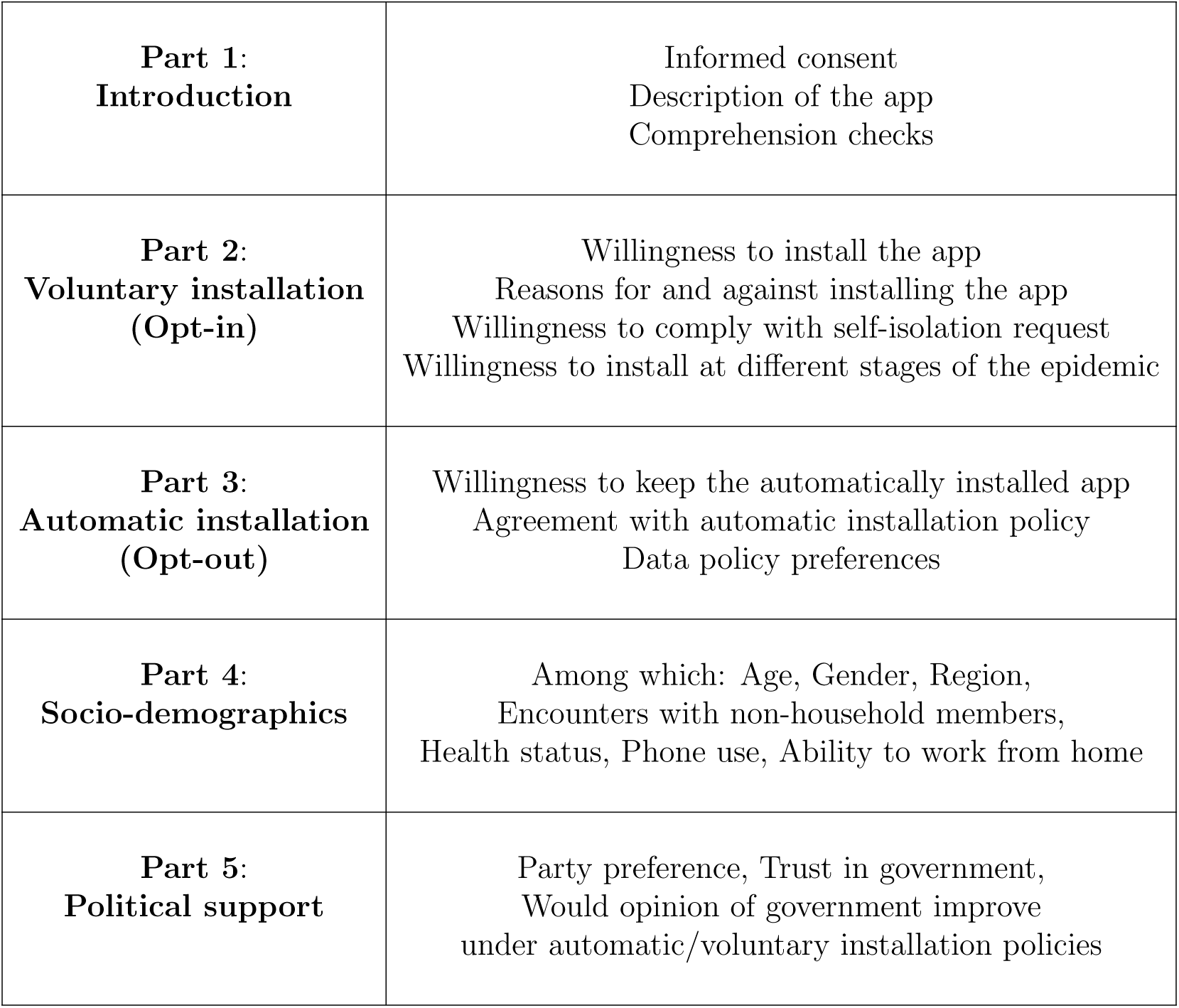
Structure of the survey

|  |  |
| --- | --- |
| **Part 1:**<br>**Introduction** | Informed consent<br>Description of the app<br>Comprehension checks |
| **Part 2:**<br>**Voluntary installation**<br>**(Opt-in)** | Willingness to install the app<br>Reasons for and against installing the app<br>Willingness to comply with self-isolation request<br>Willingness to install at different stages of the epidemic |
| **Part 3:**<br>**Automatic installation**<br>**(Opt-out)** | Willingness to keep the automatically installed app<br>Agreement with automatic installation policy<br>Data policy preferences |
| **Part 4:**<br>**Socio-demographics** | Among which: Age, Gender, Region,<br>Encounters with non-household members,<br>Health status, Phone use, Ability to work from home |
| **Part 5:**<br>**Political support** | Party preference, Trust in government,<br>Would opinion of government improve<br>under automatic/voluntary installation policies |

After this introductory part, the main questionnaire began, which was designed to address the following three objectives:

1. **Assess general support for adopting the app** (and complying with its requests);
2. **Identify the main drivers of adoption intentions** (reasons for and against);
3. **Evaluate support for different implementation policies** (opt-in vs. opt-out installation regimes).

First, respondents were asked to assess their likelihood of installing (or not) the app on their phone; responses were collected on a 5-point scale, from *Definitely install* to *Definitely won’t install*.^1^ Respondents were then asked about their main reasons for and against installing the app; in both cases, respondents could select multiple reasons from a menu of options (the order of these options was randomized at the individual level). We subsequently asked respondents how likely they would be to comply with the request to self-isolate for 14 days if they had been in close contact with an infected person. Responses were collected on a 5-point scale from *Definitely comply* to *Definitely won’t comply*. Those who did not select *Definitely comply* were then asked whether their chances of compliance would increase, decrease, or remain the same if health services committed to test them for the virus within two days from the start of their self-isolation (a negative test allowing them to stop self-isolating). Respondents who did not say they would *Definitely install* the app in response to the initial installation question were asked further questions about their willingness to install the app in three additional scenarios: (i) in case the epidemic had spread to someone in their community, (ii) to someone they knew personally, (iii) or in case an “all clear” message by the app would be associated with a relaxation in lockdown restrictions.

We next assessed whether respondents would be open to an “opt-out” policy: the government would require mobile phone providers to automatically install the app on all phones, but users would be able to immediately uninstall it. Respondents were asked about their willingness to keep (vs. uninstall) the app in this case (on a 5-point scale from *Definitely keep* to *Definitely uninstall*). As a follow-up question, they were asked to rate the extent to which they agreed with the following statement: “The government should ask mobile phone providers to automatically install the app on all phones” (on a 5-point scale from *Fully agree* to *Fully disagree*). Respondents who did not *Fully agree* with the statement were then asked whether their opinion would change if someone in their community, or someone they knew personally, had been infected with the virus. Finally, they were asked for their preference regarding the data policy that should be adopted once the epidemic is over: automatically delete all data, de-identify the data and make it available for research purposes, or some other option of their choice.

The third block of the survey collected basic demographic information (age, gender, region of residence) as well as information about potential risk factors for contracting the virus (frequency of social interactions and health risks), use of smartphone, and ability to work from home and receive (some fraction) of payment.

The final block consisted of questions about political orientation and attitudes towards the government. We first asked respondents to state their political affiliation. We then asked them whether, in general, they “trusted the government to do what is right”. Finally, we asked them whether their opinion of the government would improve in case of (i) an opt-in installation regime, and (ii) an opt-out regime.

We kept the survey design as similar as possible across all five countries, with a few exceptions. First, the UK survey was administered before the government issued a nation-wide lockdown. On the other hand, respondents in Germany, Italy and France, were surveyed *after* such a lockdown had been implemented. In the US, most states were under a “stay at home” order by the time of the survey, but not all of them. To reflect this difference in the environment respondents faced, we slightly adjusted the phrasing of some of our questions between areas in lockdown and those that were not.^2^ Second, unlike in the other countries, we asked UK and US respondents about an app that might use GPS data in addition to Bluetooth. Third, in the US survey, which was run last, we included a question explicitly asking respondents which installation regime (opt-in or opt-out) they preferred. We did this in order to check the consistency between their actual preferences for installation regimes and their installation intentions under each regime elicited in Parts 2 and 3. We also added demographic questions about ethnicity, area of residence, health insurance and media use, as well as two versions of a question about willingness to install if a private company like Facebook endorsed it. A detailed overview of the entire survey flow explaining the survey logic and highlighting differences between countries can be found in Section D in the Multimedia Appendix. The full texts of the different surveys can be found here: UK, France, Germany, Italy and US.

#### A.2 Technical Details

The survey was administered between the 20th and 27th of March 2020 in the four European countries (France, Germany, Italy, UK), and between the 7th and 10th of April 2020 in the US. No personal data was collected at any point during the survey and we obtained informed consent as well as checked for bots before the survey began. Respondents who accepted the survey invitation were directed to our online questionnaire, programmed using the software Qualtrics. The survey was pretested before fielding by generating test observations (around 4000 iterations for each country) to check data quality and consistency. Moreover, we always had a soft launch before the full launch. That is, after collecting the first 100 responses, we paused recruitment and checked the data before launching the survey widely. No adjustments had to be made in any of the countries. Our survey was not password protected but distributed through an open link, so in principle respondents could take part multiple times. However, the panel provider Lucid assigns a unique ID to each respondent which we tracked within the survey, which allowed us to identify and exclude multiple entries from the same respondent. Only a very small number of respondents completed the survey more than once; see Section B.2 for more details. Throughout the survey, respondents were allowed to navigate back and forth between screens except for when adaptive questioning was used. Furthermore, we enforced responses in all questions but those about reasons for and against installation (as we did not want to force people in favor of/opposed to installation to give reasons against/for, respectively). Consequently, every participant who submitted the survey also replied to every question. We did give people the option to choose “Don’t know” in response to all the installation questions (as well as a question about sick payment in the demographics part); however, since generally not many people chose it, we merged this option with the midpoint of the scale during data analysis. The entire survey was stretched over 17-23 screens (21-28 screens in the US), depending on the adaptive questioning, with 1-5 items per screen. The average completion time was 11.0 minutes in France, 9.7 minutes in Germany, 10.3 minutes in Italy, 8.7 minutes in the UK, and 12.6 minutes in the US.

### B Sample

#### B.1 Recruitment

In all five countries, we recruited respondents through Lucid, an online panel provider that works with a variety of sample suppliers to get a broad range of volunteers. Volunteers were recruited using a multitude of methods, from double opt-in panels (the vast majority), publishing networks, social media, and other types of online communities. In some cases, participants were furthermore recruited offline e.g., via TV and radio ads, or with mail campaigns. Participation was voluntary and, in the majority of cases, incentivized with most suppliers providing loyalty reward points or gift cards, and some providing cash payments. In each country, we set recruitment quotas so as to achieve a sample of respondents that was representative of the adult population of the respective country with regard to gender, age and region of residence (quota-based sampling). Furthermore, we did not invite individuals who did not own a mobile phone.^3^

#### B.2 Attrition and Final Sample

Figure 3 shows how many individuals opened the survey by clicking on the link as well as the subsequent attrition at different stages that left us with our final sample of 5995 complete responses. Across the five countries, 10375 individuals started the survey, out of which 10308 consented to participate (a participation rate of 99%). Before participants could begin the main part of the survey, we briefly described the app, and asked three comprehension questions to ensure participants were paying adequate attention and not just (randomly) clicking through the survey. Only if all three comprehension question were answered correctly were respondents allowed to continue with the survey. Out of the people who consented to participate, 3983 failed to answer all three comprehension questions correctly, leaving us with a sample of 6166 respondents who started the main questionnaire. 105 of these respondents either exited the survey before submitting or took the survey more than once and were therefore excluded from the sample,^4^ which left us with 6061 complete and unique responses (a completion rate of 59%). Finally, we dropped observations from 44 individuals who indicated in the comments section they did not own a smartphone, and 22 individuals who either did not identify as male or female, or who preferred not to disclose their gender.^5^ This gave us a final sample of 5995 participants for whom we have responses to all questions.

**Figure 3:**
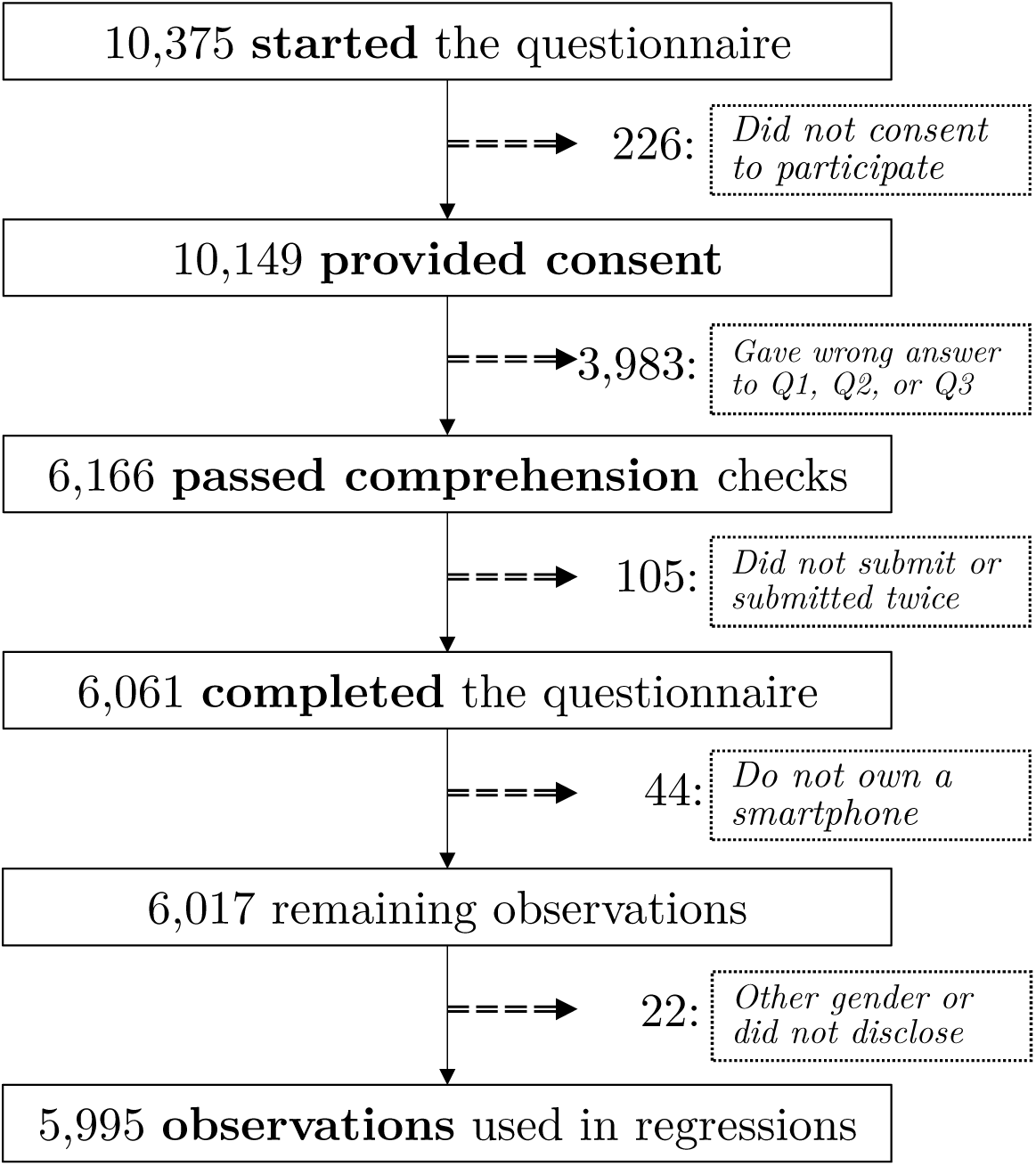
Attrition and final sample

**Table 2:**
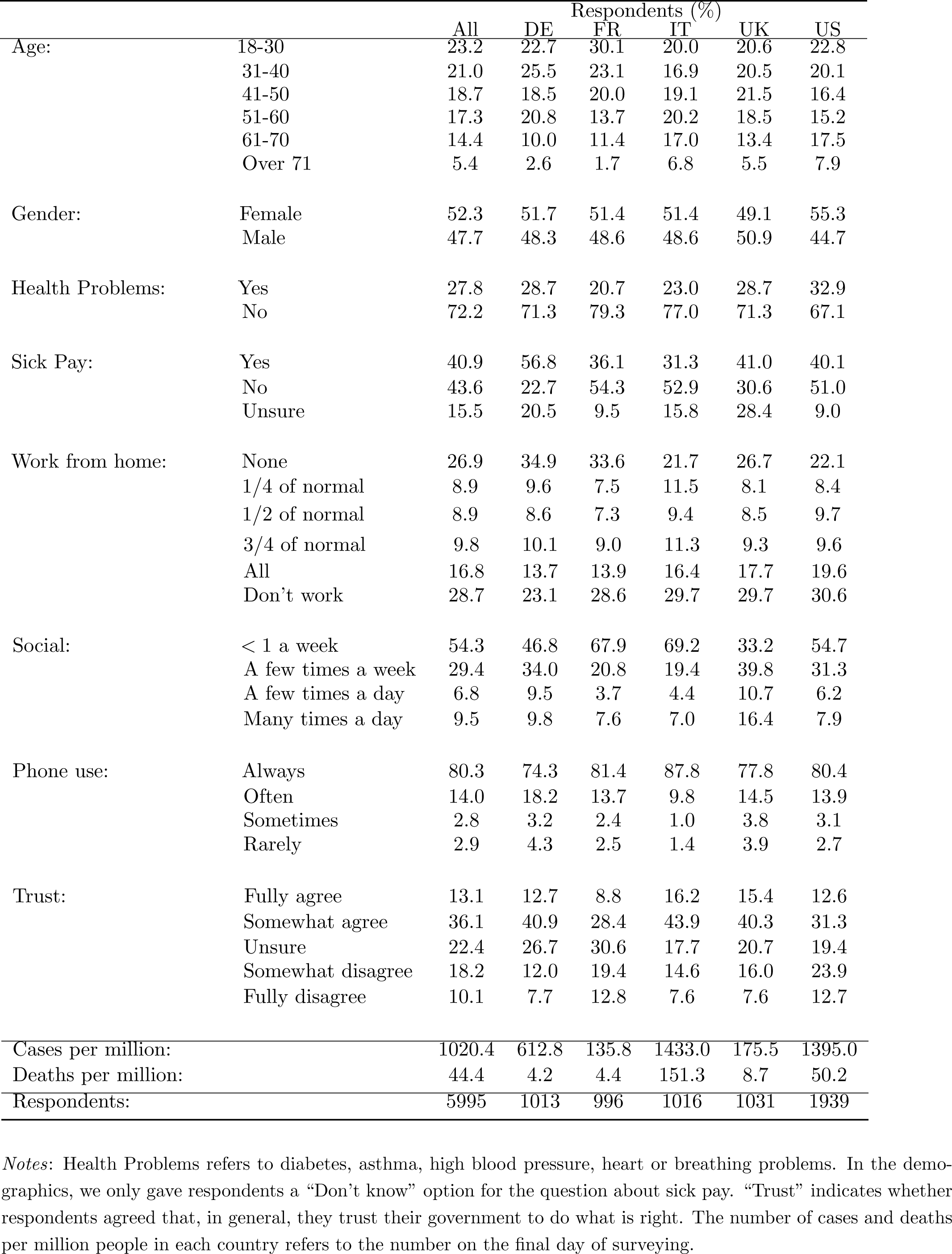
Demographics table

#### B.3 Sample robustness

Lucid recruits survey participants online through a variety of sampling partners. From these partners, Lucid receives demographic information about the pool of potential participants and uses this information to target respondents in accordance with the quotas set for the survey. In our case, we targeted the gender and age composition as well as the regions of residence of the respondents to be representative of each country’s overall population. After sending the survey out to an initial sample meeting these quotas, Lucid subsequently focuses on particular demographic groups in the re-sampling to meet the quotas in the final sample of participants (i.e., if initial take-up among people over the age of 60 was low, Lucid would disproportionately target them or even close data collection on younger cohorts completely after a while to ensure balance across age groups). However, while we tried to ensure that our sample was representative of each country’s population in terms of gender, age and region of residence, the composition could still differ from the country averages in terms of other characteristics. Furthermore, respondents who are recruited online may be more tech savvy and willing to use a phone application than the average individual. This could potentially bias our estimates.

To address these concerns and assess the external validity of our sample, we investigated its representativeness in two ways, using the German sample as an example. First, we re-created our key figures using sampling weights to harmonize the characteristics of our sample with the German population at large. Our results remained generally consistent - see for example Figure 4 which depicts the re-weighted response likelihoods to the question “How likely would you be to install the app on your phone”. However, if unobserved factors like tech savviness are not strongly correlated with demographics, then using survey weights alone is not enough to un-bias the results. As a second step, we also tested whether our results are robust to alternate recruitment methods. One week after the initial survey, we repeated our online survey twice, once again with Lucid and also with Forsa. Although both surveys were conducted online, Forsa, unlike Lucid, recruits its online panel members from a probability based, randomly selected telephone sample.^6^ This offline recruitment process ensures that technical literacy and willingness to share data should play less of a role in the selection into the Forsa sample. We find almost exactly the same results in the replication survey, alleviating concerns about our original sample – see, for example, Figure 4 which depicts results for the question “How likely would you be to install the app on your phone”. More details about the replication can be found in the German country report.

**Figure 4:**
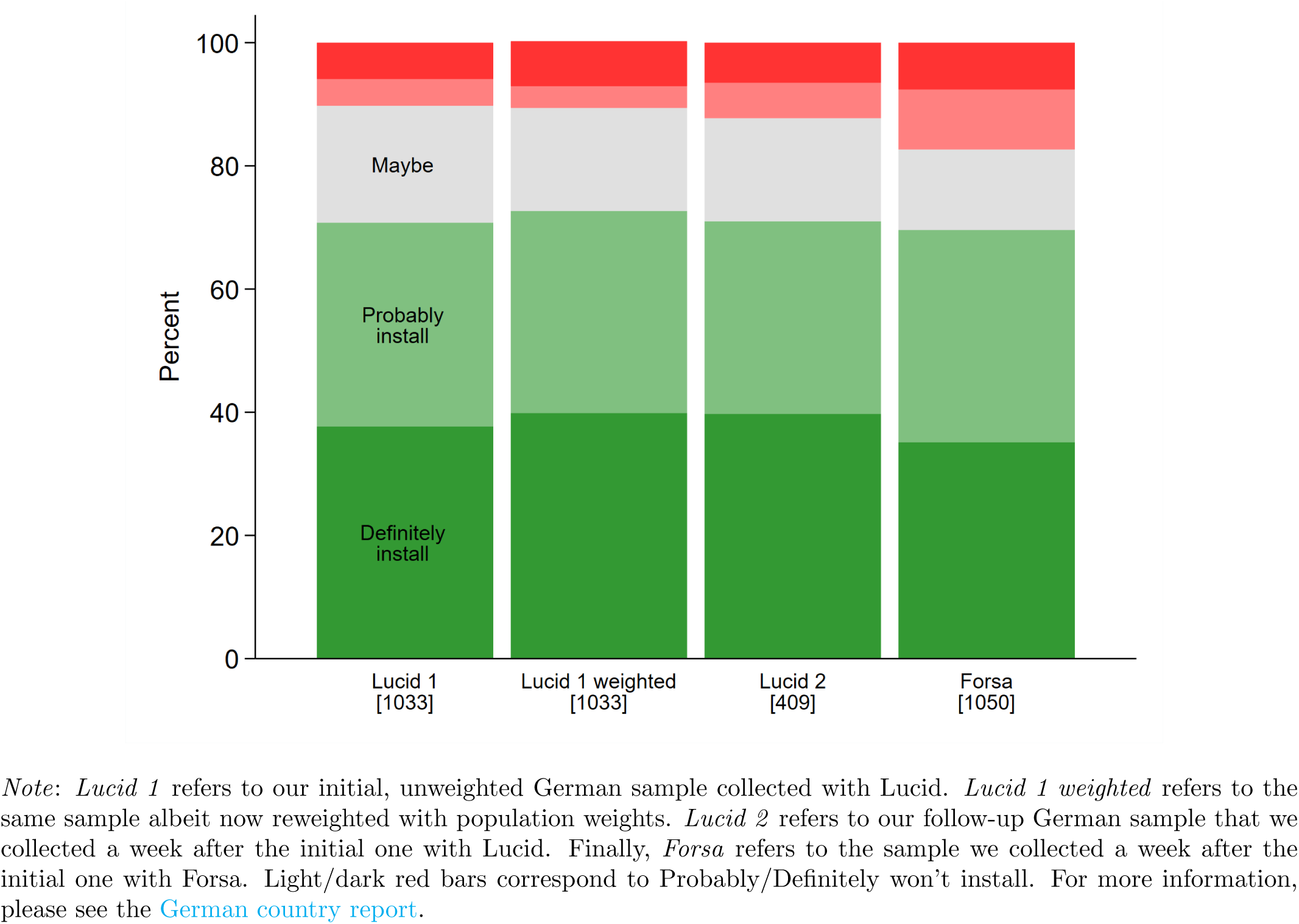
Would you install the app? – Robustness in the German sample

#### B.4 Selective Attrition

Using data from only individuals who pass all three comprehension questions may introduce selection bias in our sample. For example, more tech savvy respondents may simultaneously be more likely to pass the comprehension checks and more inclined to install any app. To address this concern, we consider a selection model where the likelihood of a respondent consenting to the survey and correctly answering all comprehension questions is a function of the demographic information we obtained from Lucid for everyone who started the survey (age, gender, region, employment status, and, in the case of Italy, France and Germany, education). We estimate the following probit model:

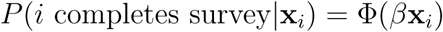

where Φ() gives the normal cumulative distribution function. We also estimate an ordered probit model, taking the stage of a respondent’s drop out (consent, comprehension question 1, 2 or 3) as our ordinal outcome variable. The results of these empirical specifications are given below. Respondents with less than high school education are significantly less likely to be included in our final sample. The same is true for male respondents, younger respondents, and the self-employed. Given our selection model, we re-weight our analyses with inverse probability weights reflecting the likelihood that each individual was sampled. Therefore, using our selection model, weights are constructed using the predicted probability of completing the survey, 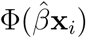. The results from re-weighting the responses to the main questions on voluntary and automatic installation are given in Figure 6. Importantly, we see that the results are broadly consistent with the results presented in the main text.

**Figure 5:**
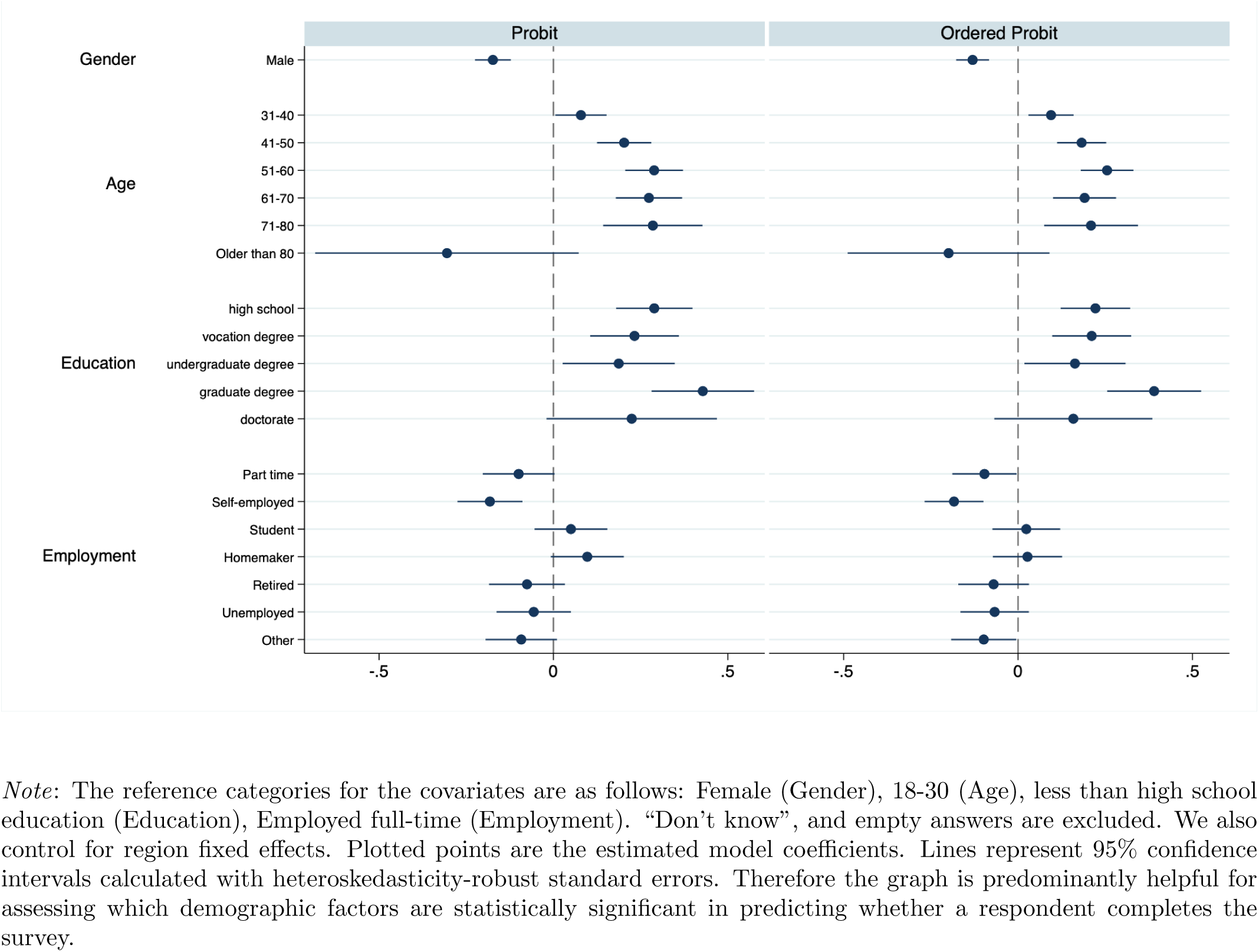
Selection model

**Figure 6:**
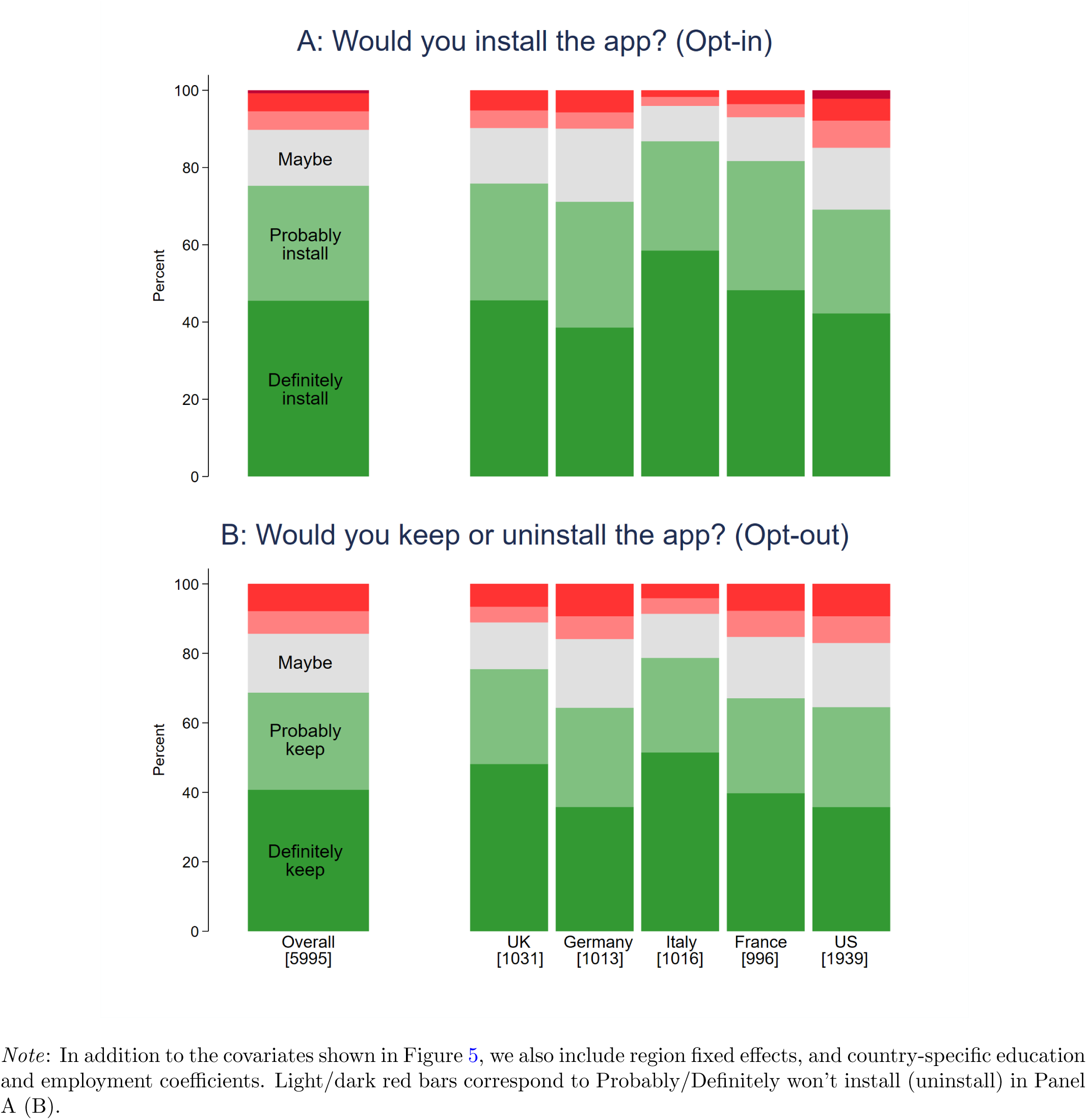
Likelihood of having the app in an opt-in and opt-out regime, weighted by inverse probability of staying in sample

Finally, we also compared the demographic information we received from Lucid to the self-reported demographic information on gender and age. This allows us to check that respondents gave accurate information in our survey. 335 respondents (5.5%) gave at least one inconsistent answer. Removing these individuals has no effect on the results.

### C Additional results

#### C.1 Robustness to modelling assumptions

In the main part, we make a couple of modelling assumptions. Namely we use a Linear Probability Model and dichotomize the outcome measure by grouping everyone who said they would *probably or definitely install* the app into the “install” category (assigned value 1) while everyone else falls into the “non-install” category (with value 0). However, our results are robust to making different modelling assumptions as well. Figure 7 displays the results of the opt-in installation question if we model the entire choice space as an ordered logit where high numbers indicate a lower probability of installing the app, so that negative coefficients here have a similar interpretation to positive coefficients in the linear probability model in the main part. We see that the results look very similar to the ones obtained with the main specification (see Figure 2).

**Figure 7:**
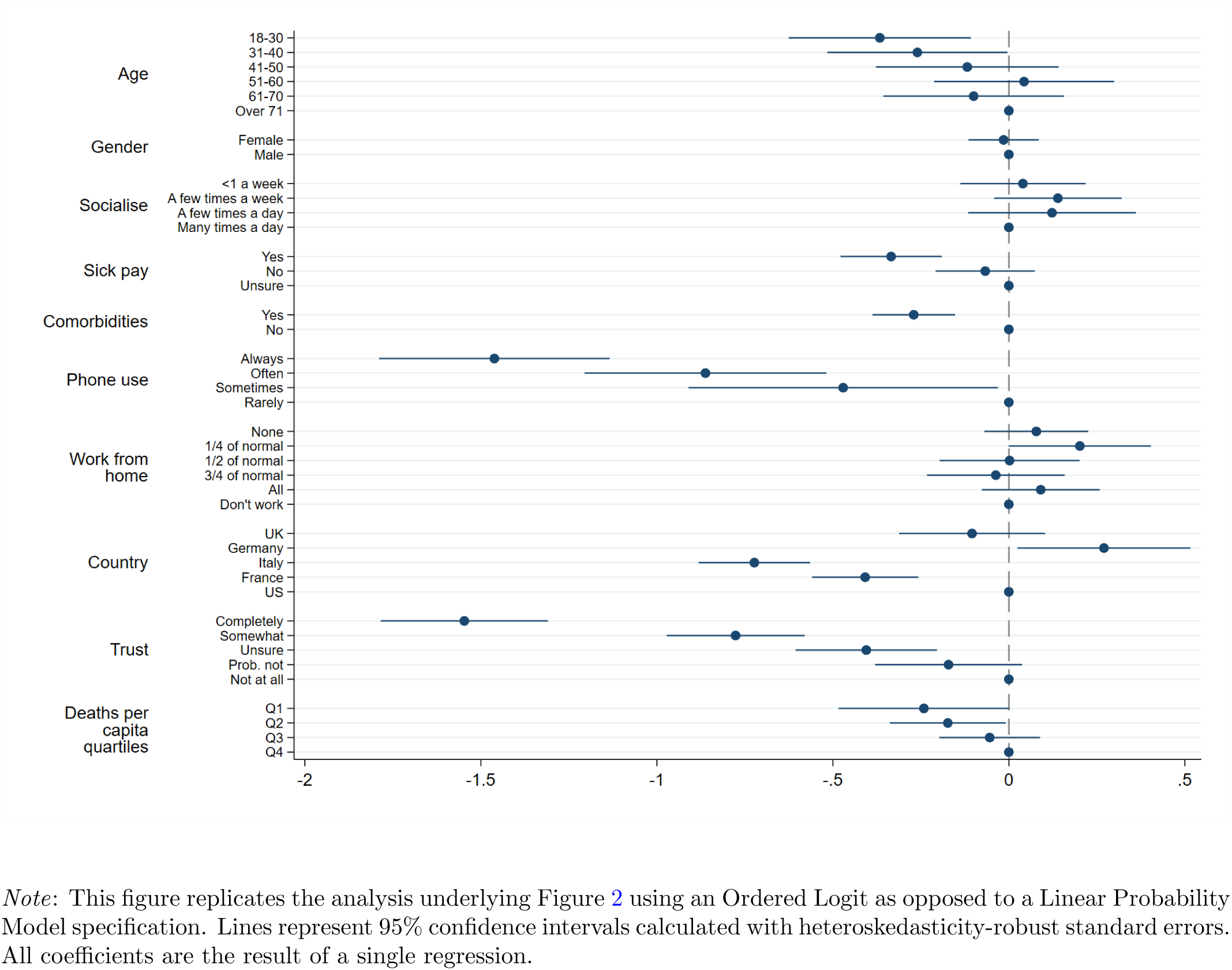
Ordered logit specification

Furthermore, our results also hold when we sort only the people who say they would *definitely* (rather than definitely or probably) install the app into the install category – both using a linear probability and a probit specification. For brevity, we do not show these results here.

Just as we have looked at the covariates determining the choice to either install or not the app voluntarily (opt-in) in Figure 2, we can also look at the impact of different influencing factors on the decision to keep or immediately uninstall an automatically installed app (opt-out). Figure 8 displays the results if we once again dichotomize the outcome so that the dependent variable takes the value 1 if a respondent indicated that they would *probably or definitely keep* the app and 0 otherwise. We see that participants from the UK are relatively more likely to keep the app in this scenario while French respondents are less likely in comparison. As with the opt-in installation outcomes, the results are robust to using an ordered logit specification as well as using a probit or linear probability specification when the “keep category” only consists of people saying they would *definitely keep* the app. For brevity, these robustness checks are omitted here.

**Figure 8:**
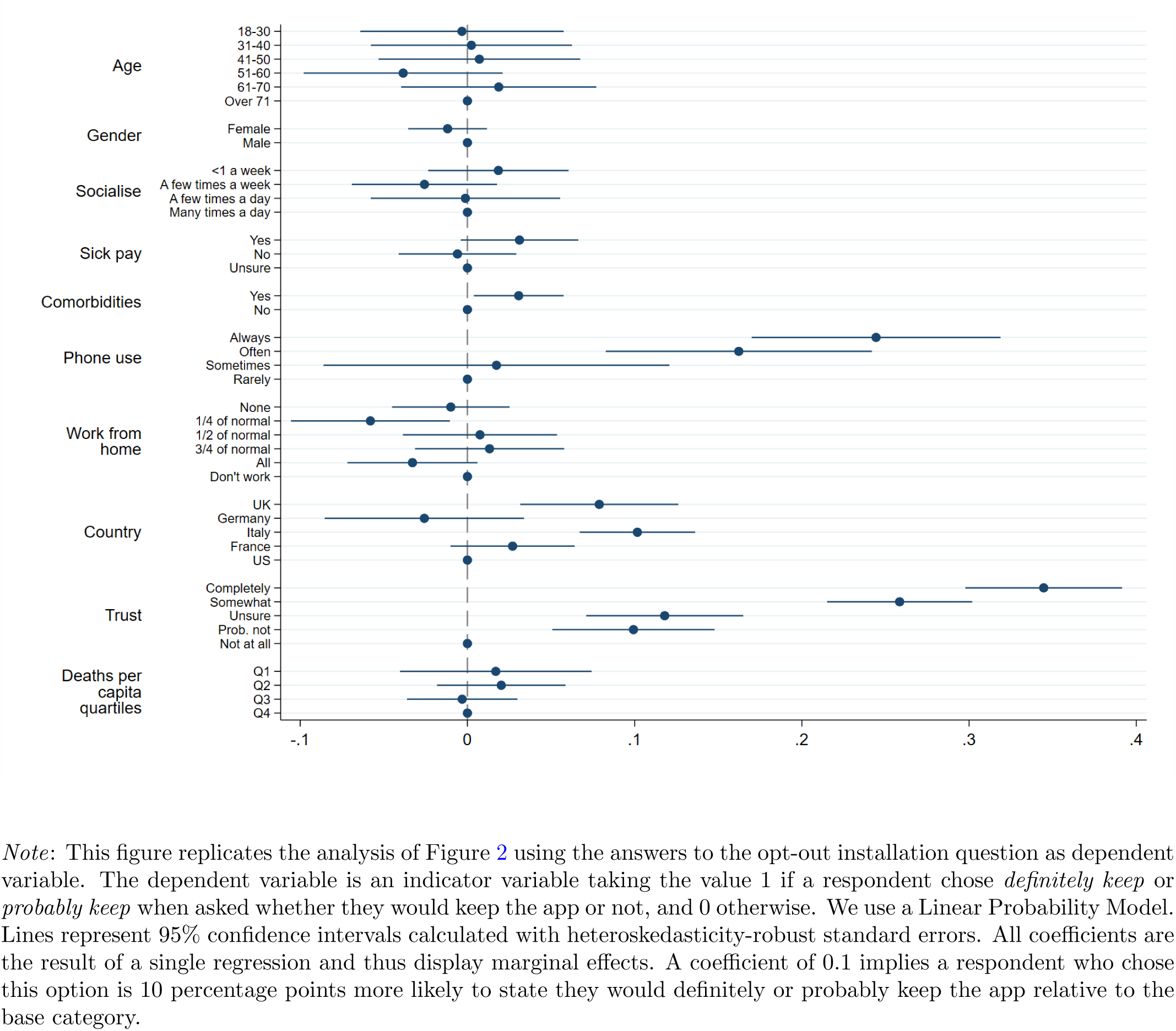
Determinants of stating *definitely or probably keep*

#### C.2 Robustness across demographics

In the main text, we note that the results presented in Figure 1 hold across a variety of different demographic groups - this is illustrated in the following. Figure 9 shows that support for the app does not vary significantly when splitting outcomes by gender, the existence of comorbidities or the availability of sick pay. Figure 10 shows that there are only very small differences with regard to age. The only dimension where we see significant differences in support for the app is trust in government. Figure 11 shows that someone’s propensity to install a contact-tracing app decreases the less they generally trust the government to “do what is right”. The results of the opt-out scenario show the exact same pattern as the opt-in results. For brevity, we do not show them here.

**Figure 9:**
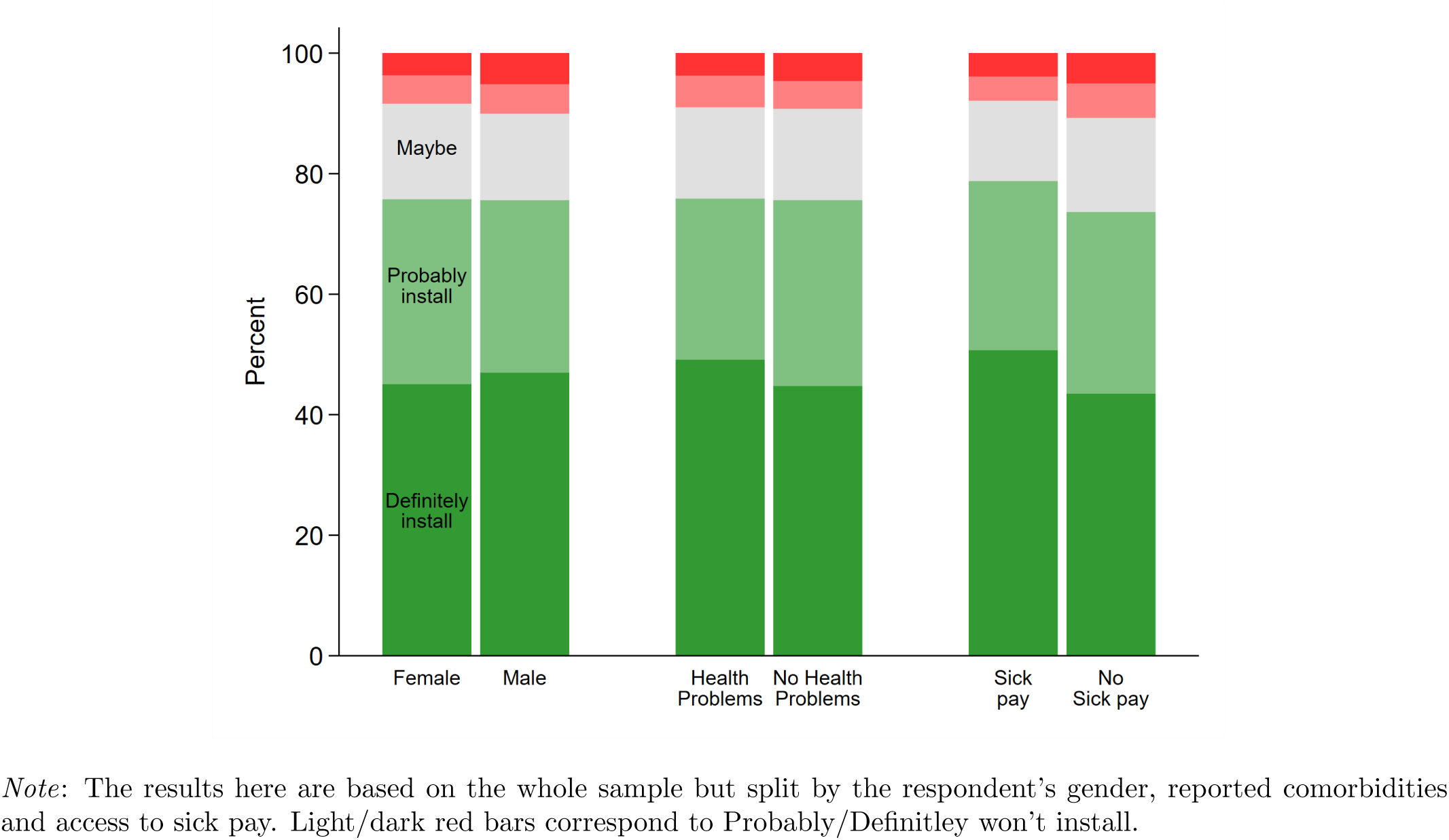
Would you install the app? (opt-in)

**Figure 10:**
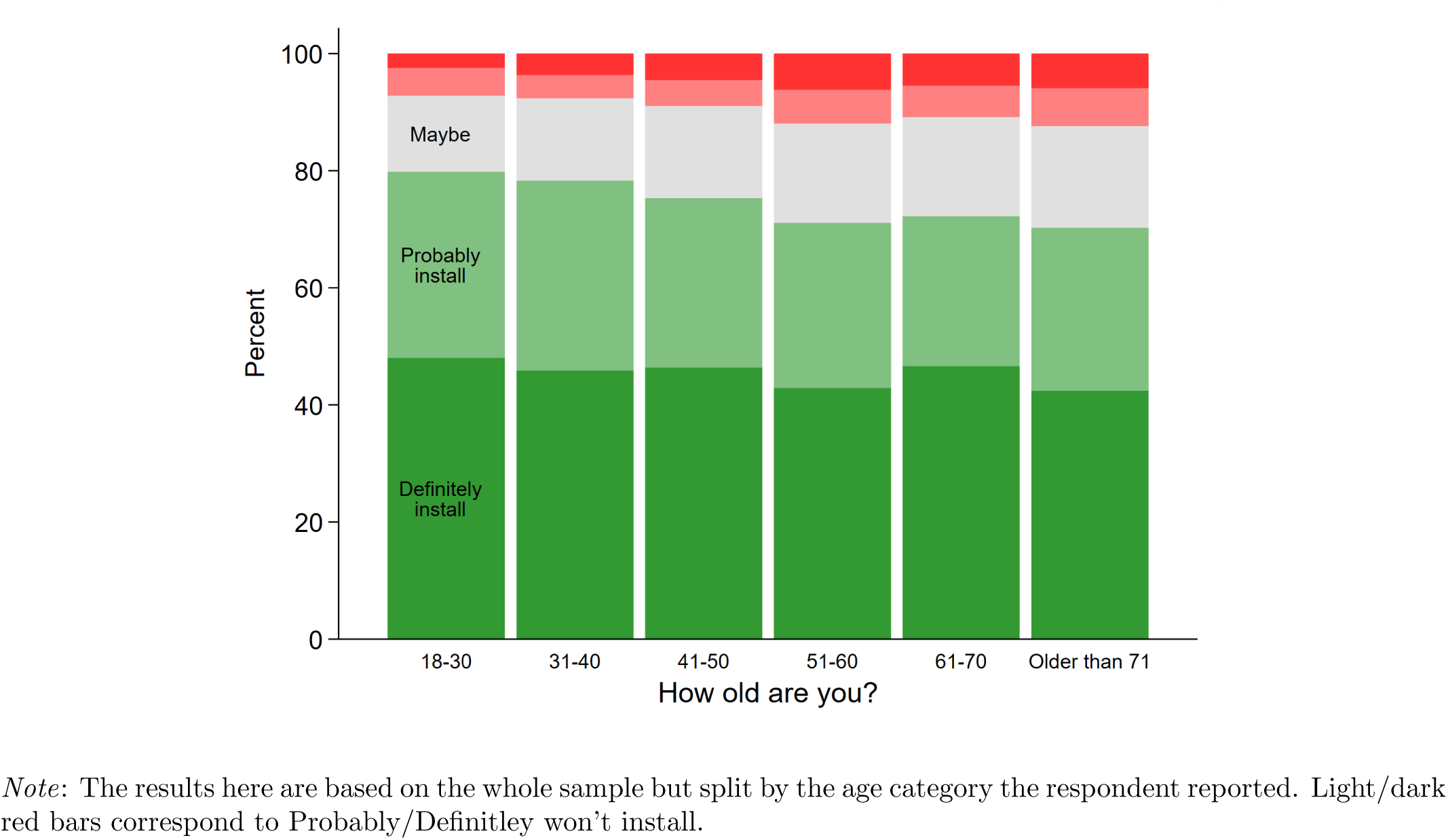
Would you install the app? (opt-in)

**Figure 11:**
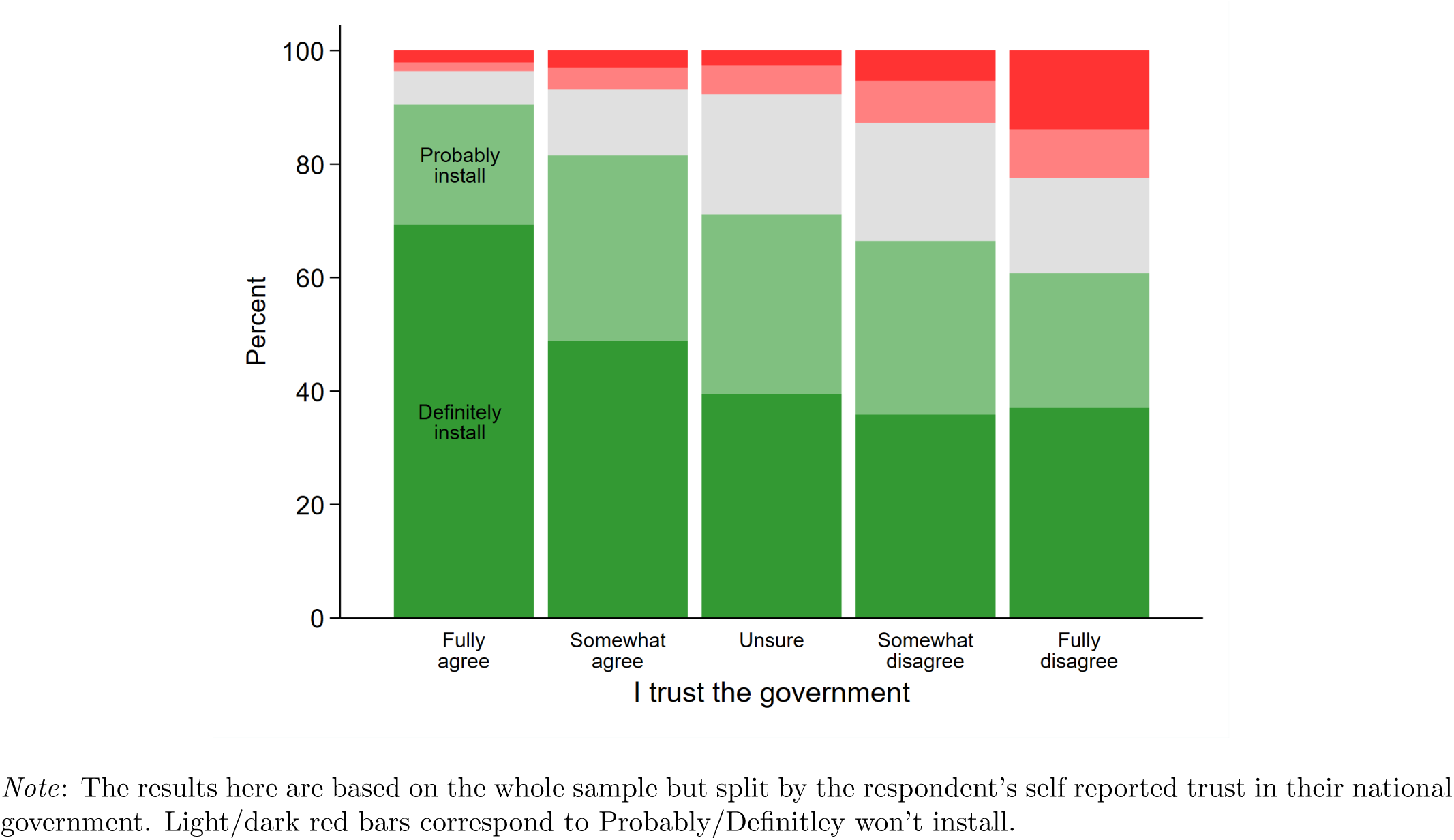
Would you install the app? (opt-in)

#### C.3 Take up and disease severity at the geographical level

We collect data on the number of infections as well as deaths due to COVID-19 at the *geographic area by day* level where the geographic area is at the same level of disaggregation as the respective survey asked for (e.g. state for the US). The data is publicly available on GitHub, and the sources are Santé Publique France (for France), Protezione Civile (for Italy), the Robert Koch Institute (for Germany), PHE (for the UK), and the New York Times (for the US). We adapted the aggregation level of the French data from *département* to region.

As can be seen in Figure 2 in the main part, we generally find only a very weak relationship between the severity of the outbreak (measured by deaths per capita as well as the absolute number of deaths) in someone’s area of residence and the probability that they will want to download or keep the app. The exception to this rule are the very badly affected areas of New York and Northern Italy – here, respondents are about 10 percentage points more likely to state they would *definitely install* the app than the average participant. The fact that we do not find a strong effect generally could be due to a number of different factors. Firstly, death rates do not vary much across geographic areas once we remove New York and Northern Italy. Secondly, much of the rhetoric surrounding the severity of the disease is at the national, not sub-national level (again excluding New York and Northern Italy). Lastly, it is likely that the cartography of infections does not exactly follow the cartography of regions at our disposal.

#### C.4 Reasons for/against installation and take up decisions

Figure 12 shows that in general, far more reasons are given in favor of installing the app than against it – this is true even among those who state that they *probably or definitely won’t install* the app. The number of reasons *for* installing the app decreases sharply as respondents become less likely to want to install it. This is not the case for reasons *against*, where most respondents select only one reason regardless of their intention to install. These findings suggest the need to better explain the various ways in which the app might benefit a user and his or her surroundings.

**Figure 12:**
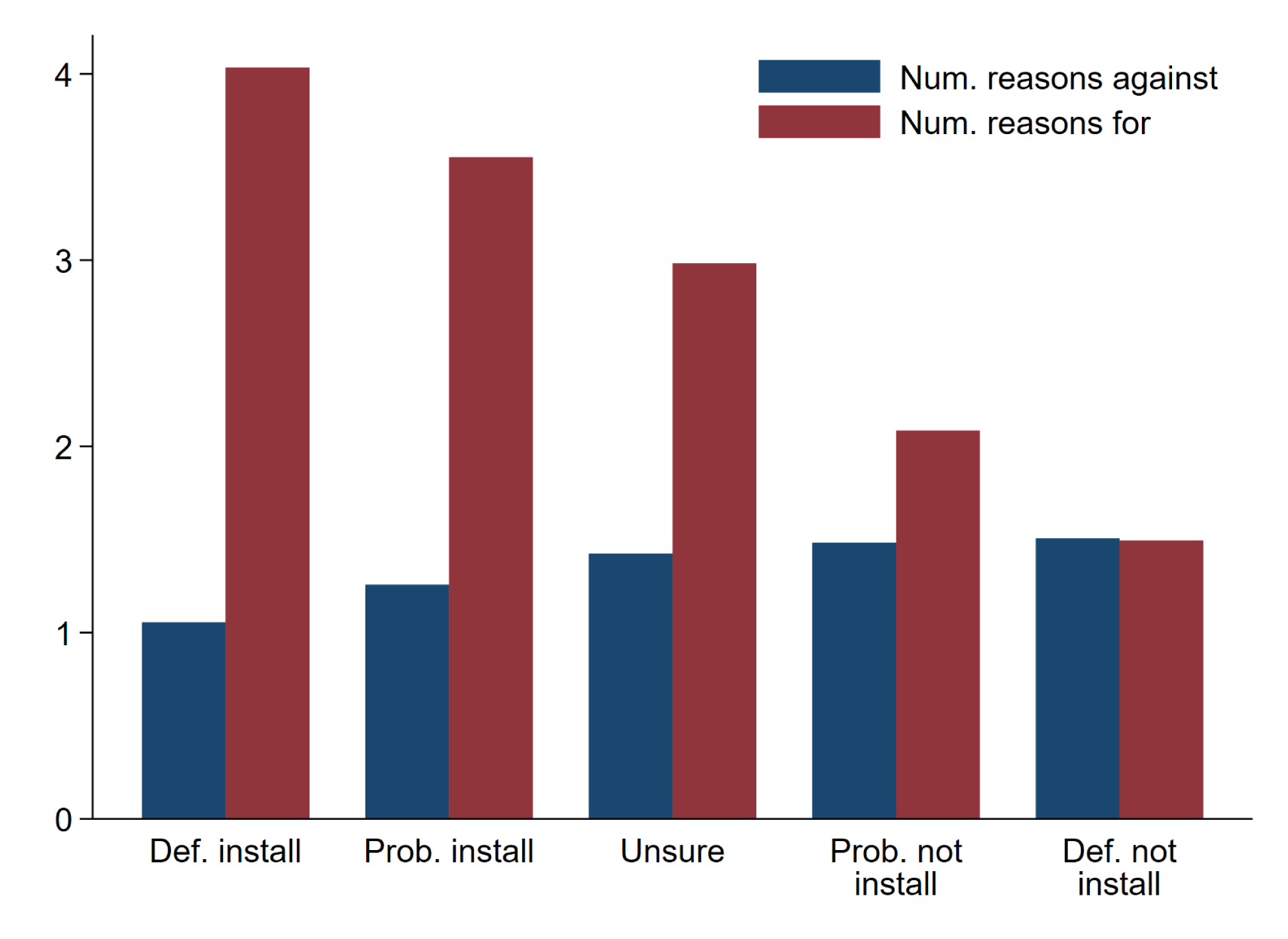
Number of reasons given for/against the app, by installation preference

The reasons chosen most often by respondents across all countries in favor of and against installing the app are shown in Figures 13 and 14. While all reasons in favor of an installation appear to come somewhat evenly to people’s minds, there is a strong clustering of reasons against the app around surveillance and security concerns as well as possible impacts on mental health. These reasons highlight the biggest concerns that would need to be addressed in the design and implementation of the app to make sure take-up would be sufficiently high. In addition to the common reasons displayed in Figures 13 and 14, we also gave a few answer options, which differed by country. In the UK, respondents were given “*Don’t want the NHS to have access to my location data*” as an additional reason against, which was replaced by *“I don’t want to activate Bluetooth”* in the other surveys. Germans in particular, were quite hesitant to activate Bluetooth, making it the third most popular reason against installation there. In the French, German, Italian and US surveys, respondents were given “ *Would allow me to return to normal life faster*” as an additional reason in favor – however, it was not chosen by many people. Finally in the US survey the additional reason against *“No one else will use the app*” was the second most popular reason, given by 40% of respondents.

**Figure 13:**
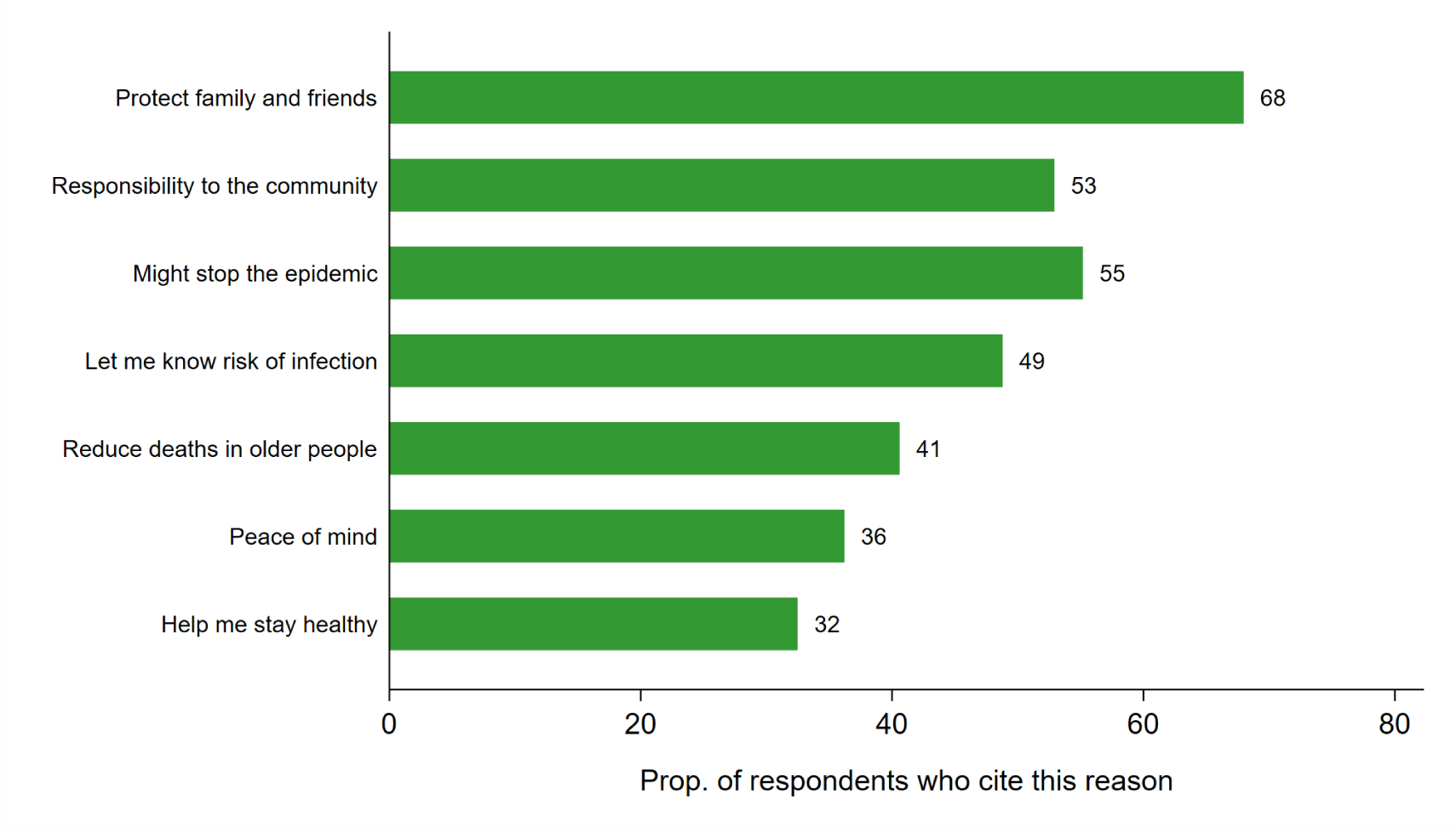
Percentage of respondents who choose each reason *for* installation

**Figure 14:**
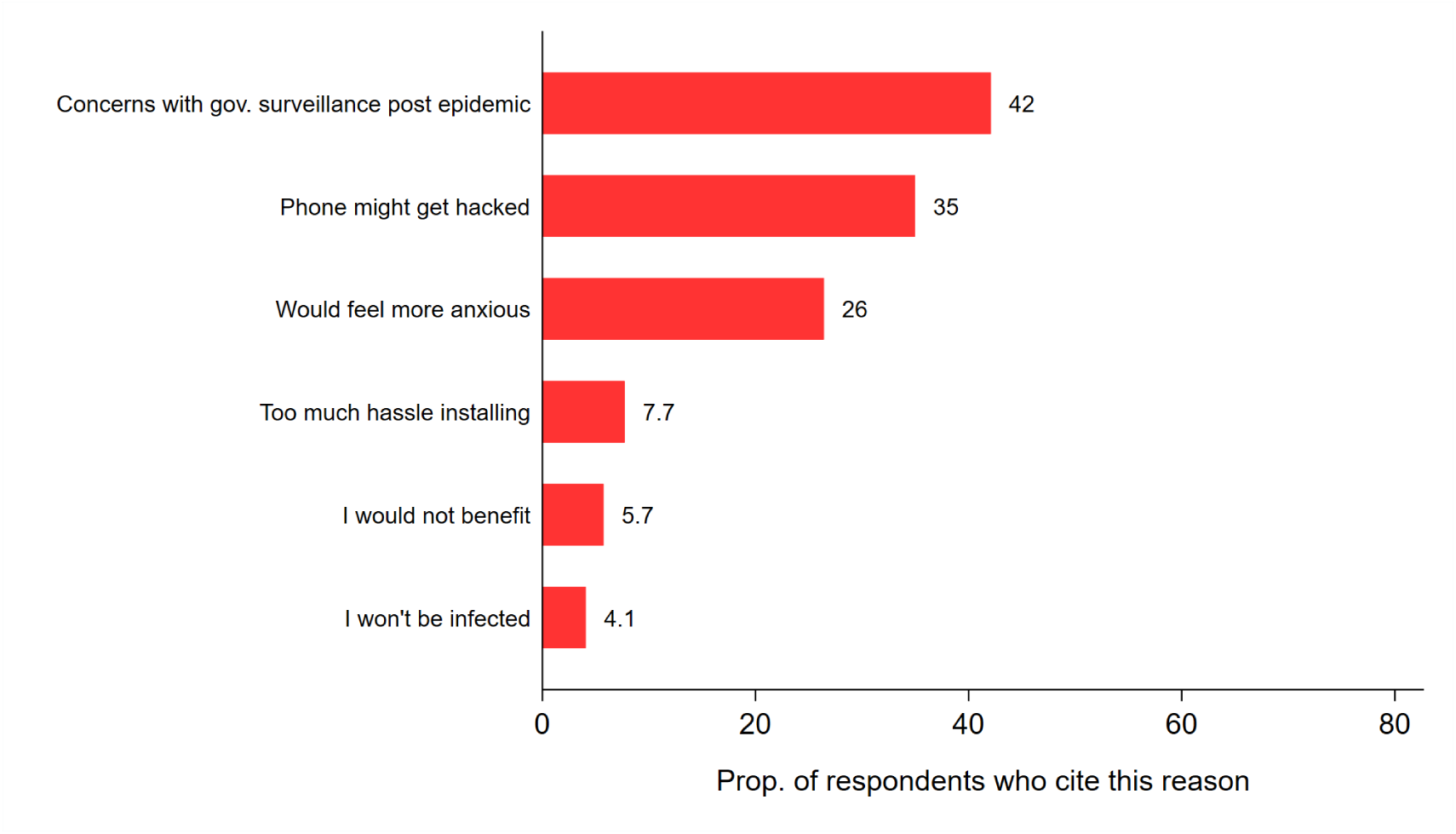
Percentage of respondents who choose each reason *against* installation

**Figure 15:**
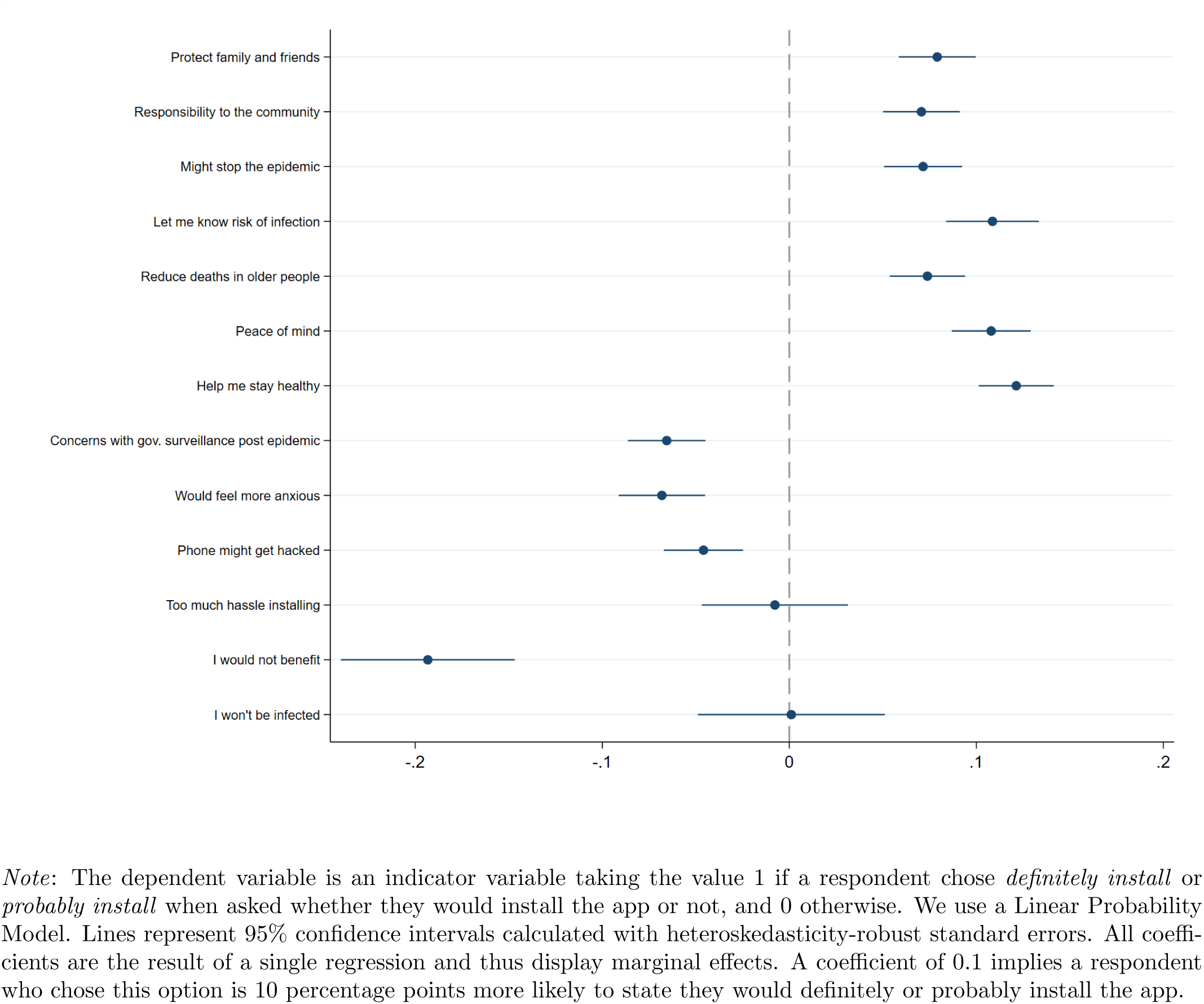
Impact of reasons on installation probability

Figure 15 shows the impact each reason had on the probability with which a respondent stated they would probably or definitely install the app – controlling for all the covariates displayed in Figure 2. Unsurprisingly, the first seven reasons, which are in favor of the app, increase the probability of intending to install the app. Strikingly, out of the six reasons listed against installation (the latter half in the graph), only one had a large impact on installation decisions and that was “I would not benefit”. This may indicate that while many people are concerned about government surveillance and cyber security, these reasons are not prohibitive of installing the app or only become so if they lead participants to conclude that the app would not be useful to them. We found similar results with an ordered logit model, a linear probability model dichotomizing on just *definitely install*, when using a probit model and dropping the controls. Finally, the results are also qualitatively similar when we look at *opt-out* rather than opt-in decisions.

#### C.5 Determinants of the reasons chosen

Figures 16 and 17 display the main determinants of reasons given for or against installing the app. Each column represents a separate linear probability model where the dependent variable takes the value 1 for individuals that chose that specific reason and the value 0 otherwise. The main takeaway from the graphs is that the reasons are largely not explained by the covariates. Only people who trust the government less as well as younger people are significantly more likely to name concerns surrounding post-epidemic surveillance as (one of) their main reasons against installation. Furthermore, younger people are less likely to cite *peace of mind* as a reason for installation. All results hold when using a probit model as well.

**Figure 16:**
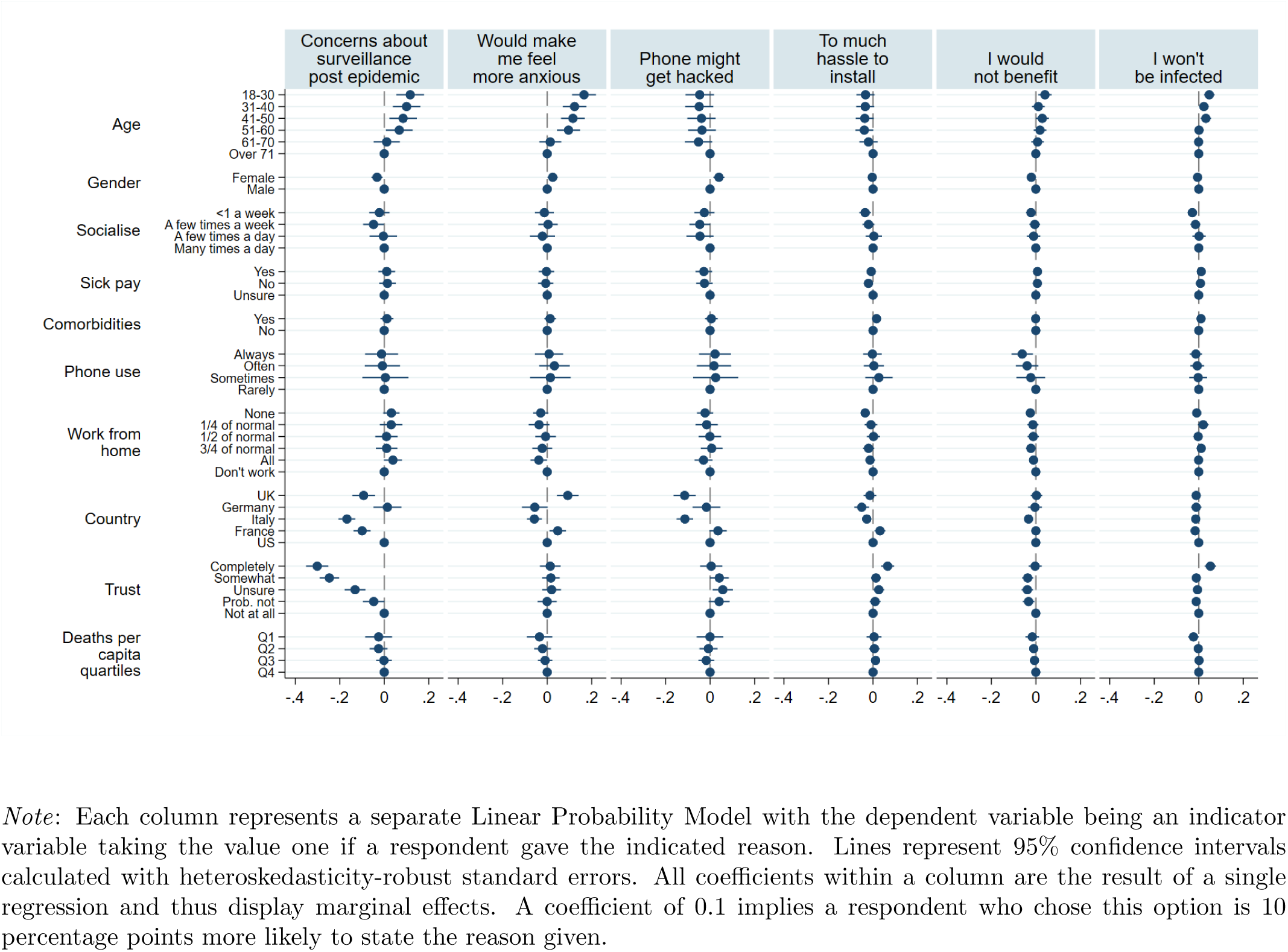
Determinants of reasons given against installation

**Figure 17:**
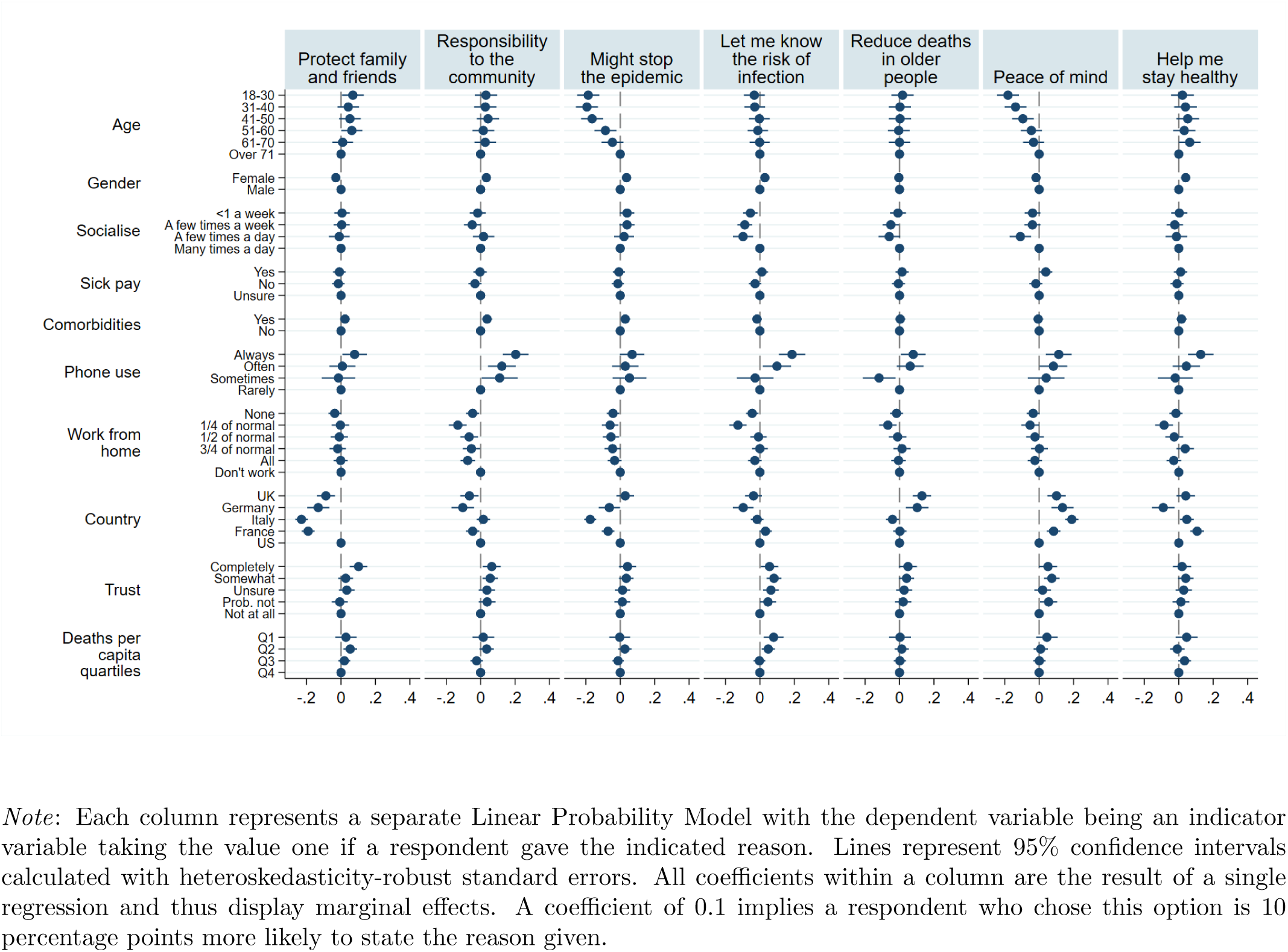
Determinants of reasons given for installation

#### C.6 Opt-in versus Opt-out preferences

Understanding whether an opt-in or an opt-out regime will yield a larger number of app users is crucial for the successful implementation of app-based contact tracing. Standard results from psychology suggest that many more people will keep the app in an opt-out regime than would install it voluntarily in an opt-in regime, as the former approach would reduce (mental) transaction costs and implicitly set having the app as the societal standard. However, Figure 1 shows that instead fewer people indicate they would keep the app rather than voluntarily download it. One reason for this may be that individuals, and in particular individuals who are concerned about potential government surveillance, may perceive automatic installation as an overreach by the government and thus choose to uninstall the app on principle.

To further understand respondents’ preferences over an opt-in vs. opt-out regime, we directly asked US respondents which regime they would prefer. 60% would prefer voluntary to automatic installation. This fraction is constant across gender, region, political affiliation, lockdown status and other characteristics.

We can infer the preference between voluntary or automatic installation regime indirectly for the other four countries, where we did not ask directly for this preference. We use participants’ intentions to install or keep the app in an op-in vs. opt-out regime to create an ‘Intention measure’. We use participants’ opinion of the national government in response to it introducing either regime, to create an ‘Opinion measure’.

The Opinion measure considers respondents’ answers to the questions:

- How much do you agree with the following statement? “My opinion of the government would improve if they introduced the app, and allowed me to decide whether to install it or not”
- How much do you agree with the following statement? “My opinion of the government would improve if they asked mobile phone providers to automatically install the app, and allowed me to decide whether to keep it or not”

The difference between respondents’ answers to these two questions yields information about which regime they would prefer the government to introduce. Therefore, we create a variable that gives the difference between these two answers, such that when participants’ opinion of the government would improve more (or worsen less) under the opt-in regime, this difference is negative. In this case, we would infer that they prefer an opt-in regime.

The Intention measure considers respondents’ answers to the “Would you install this app?”, and “Would you keep this app if it was automatically installed” questions. The difference between respondents’ answers to these questions reflects their different intentions of having the app on their phone in either regime. Thus, this difference is again indicative of participants’ preferences over the opt-in/opt-out regimes. As for the Opinion measure, when the Intention measure is negative we can infer that a respondent believes they would be more likely to have the app installed on their phone in an opt-in regime, which we take to mean that they prefer the opt-in regime.^7^

For both difference measures, we re-code the new variable into three categories: *d <* 0 (prefer opt-in), *d* = 0 (indifferent), and *d* > 0 (prefer opt-out). The differences in preferences, across countries, are displayed in Figure 18. The trends are broadly in-keeping with the other results.

**Figure 18:**
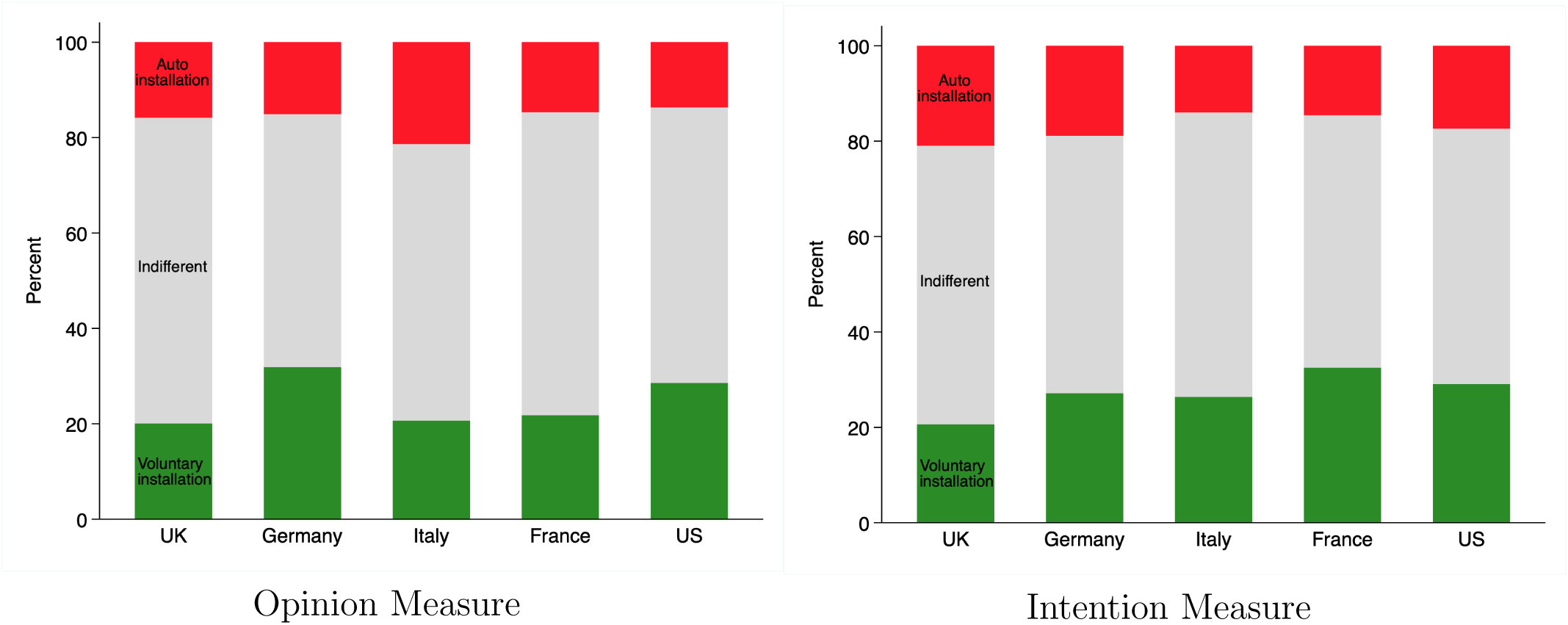
Opt-in/opt-out preferences in each country

To investigate the factors that are associated with opt-in/opt-out preferences, we use a simple ordered logit model. The results from this analysis are presented in Figure 19. Respondents who express less trust in the government or who worry about government surveillance or their phone being hacked, are more likely to have a preference for the opt-in over the opt-out regime.

**Figure 19:**
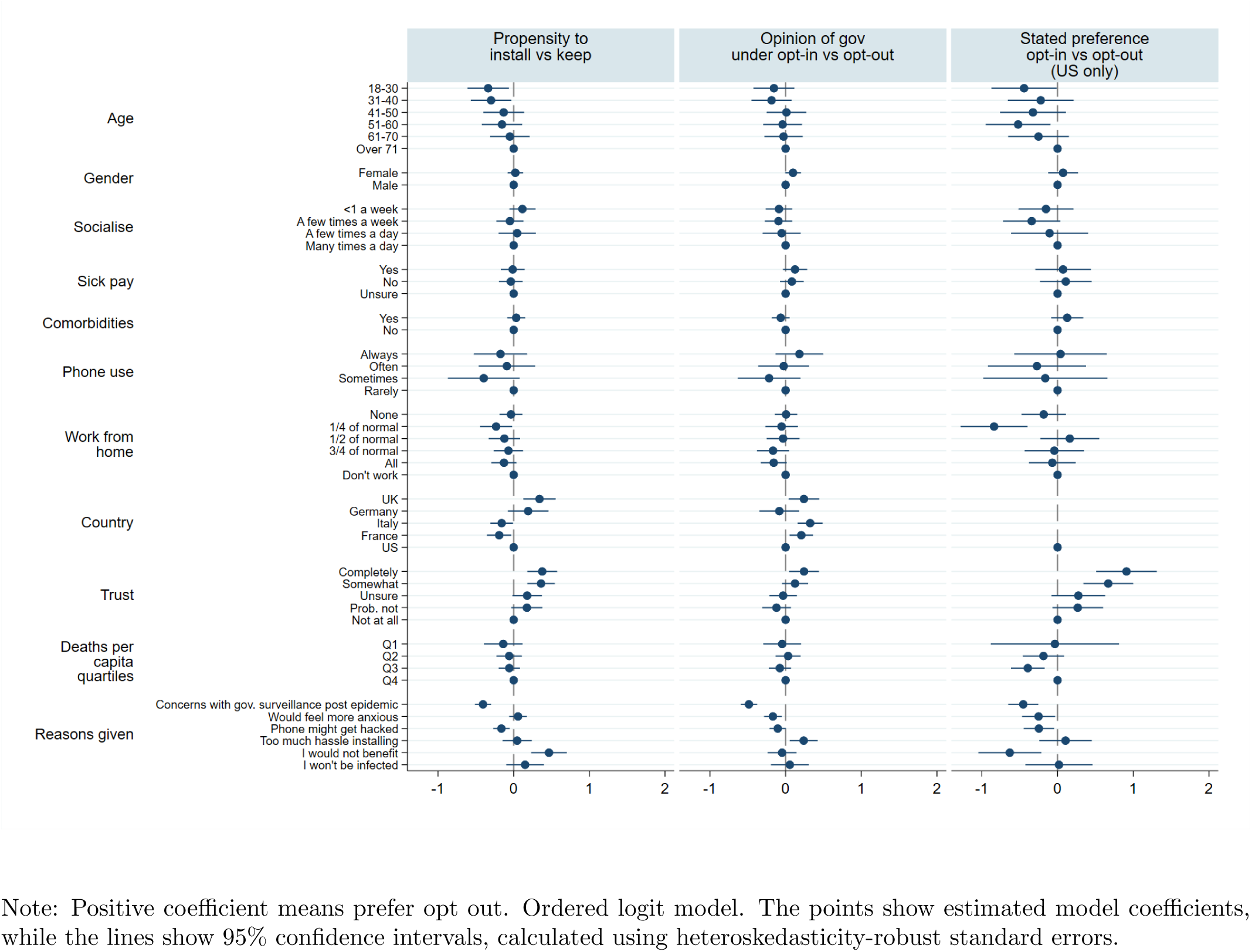
Determinants of opt-in vs. opt-out preference

##### C.6.1 Validity of difference measures

In the US survey, respondents were asked directly whether they would prefer an opt-in or opt-out regime. The below cross tabulation demonstrates a strong relationship between respondents’ stated choice (columns) and their inferred preference (rows), using the Opinion Measure. Only 10.4% of respondents’ inferred preferences are inconsistent with their stated preferences - those who selected *d* < 0 and Automatic, or *d* > 0 and Voluntary. Of the respondents for which the Opinion measure is negative (*d* < 0), 80.4% stated that they prefer the Voluntary regime. Likewise, of the respondents for which the Opinion measure is positive (*d* > 0), 64.7% stated that they prefer the Automatic regime.

|  | US respondents |  |
| --- | --- | --- |
|  | Voluntary | Automatic |
| $d < 0$ | 445 | 109 |
| $d = 0$ | 564 | 555 |
| $d > 0$ | 94 | 171 |

#### C.7 Data Usage

Respondents were asked for their preferences over what should happen to the data generated by the app. Respondents could state that they wanted the data immediately deleted, or that the (deidentified) data should be made available to researchers.

In the final sample, 59.9% of respondents answered that the de-identified data should be made available to researchers. Figure 20 demonstrates that respondents who were less likely to say they would download the app were also less likely to want their data to be made available to researchers. Surprisingly, Figure 20 demonstrates no relationship between trust in the government and data-sharing preferences. However, once we control for other covariates, this relationship does become statistically significant, as can be seen in Figure 21. This relationship is nevertheless non-monotonic and hence difficult to interpret. Figure 21 also demonstrates that older people, people who use their phones more regularly, and residents of the UK are more likely to consent to having their data shared.

**Figure 20:**
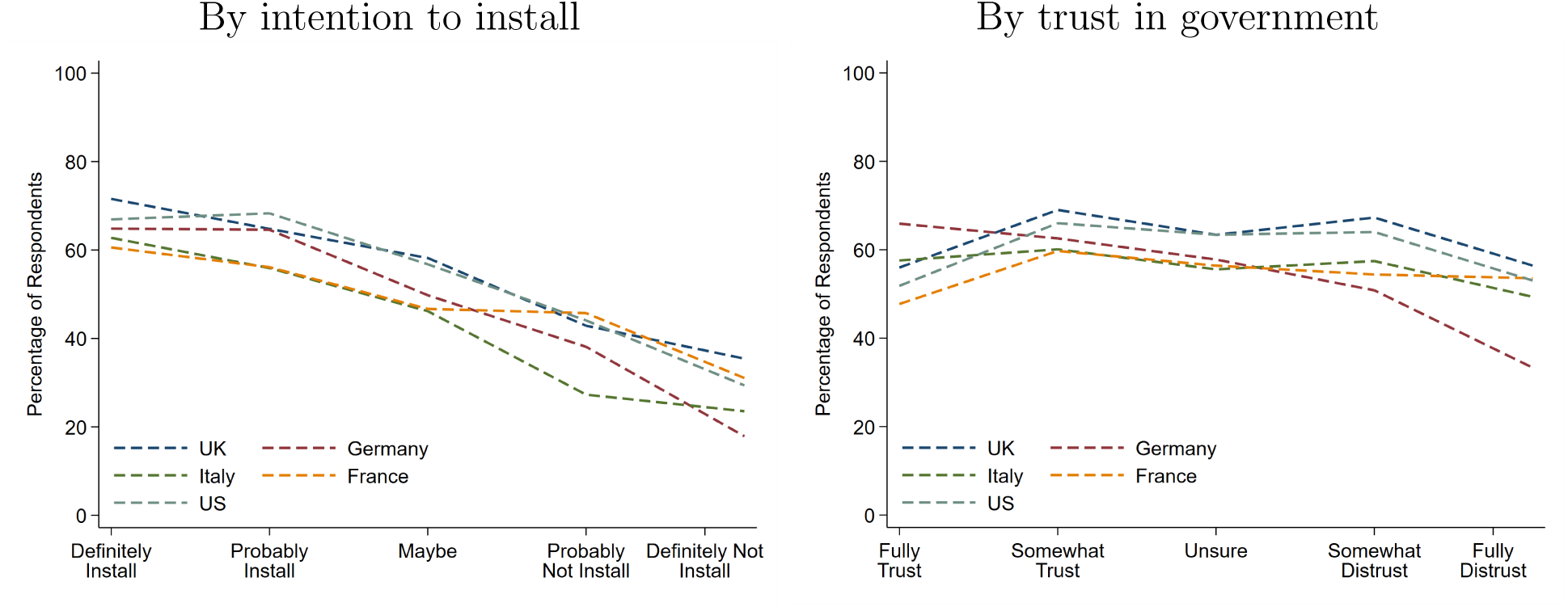
Percentage of respondents wanting the data to be made available

**Figure 21:**
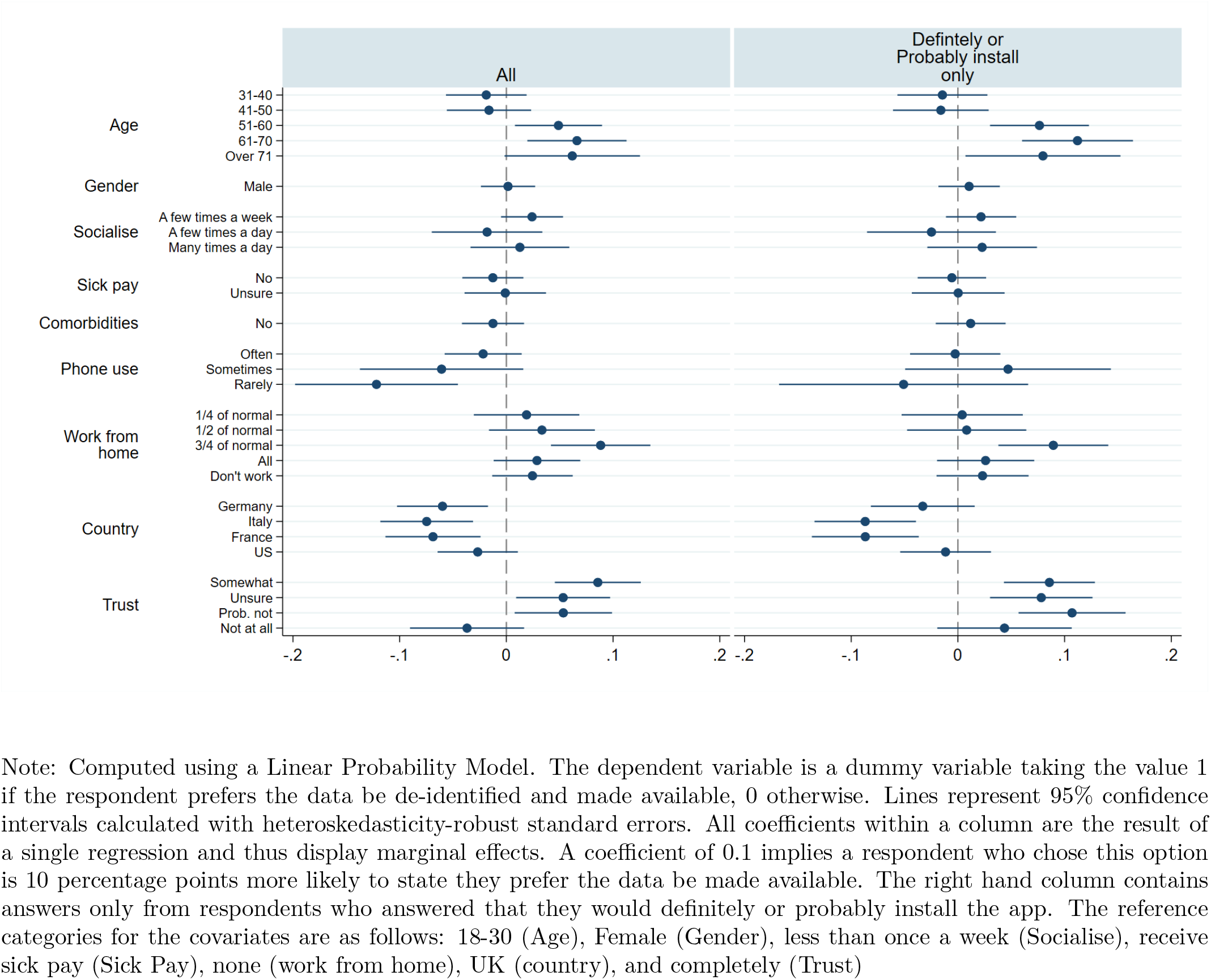
Determinants of wanting the data to be made available

#### C.8 Compliance with the app self-isolation request

We asked respondents how likely they would be to comply with the request of self-isolating for 14 days if they had been in close contact with a person who was confirmed to be infected. Responses were collected on a 5-item scale ranging from *Definitely comply* to *Definitely won’t comply*. As shown in Figure 22, the vast majority of respondents in all countries said they would comply with the self-isolation request. Support is again highest in Italy, where 96% of respondents declared they would *definitely* or *probably comply*, and lowest in Germany, at a still very high 89%. We further asked respondents who did not say they would definitely comply, whether their chances of compliance would increase, decrease, or remain the same if the health services in their own country committed to test them quickly. In all countries, a commitment to quick testing would further increase compliance rates. This implies that the vast majority of people in all five countries are not only prepared to have the app installed on their phones, but also to use it as intended, even if this means sacrificing (more of) their personal freedom for a limited amount of time.^8^

**Figure 22.**
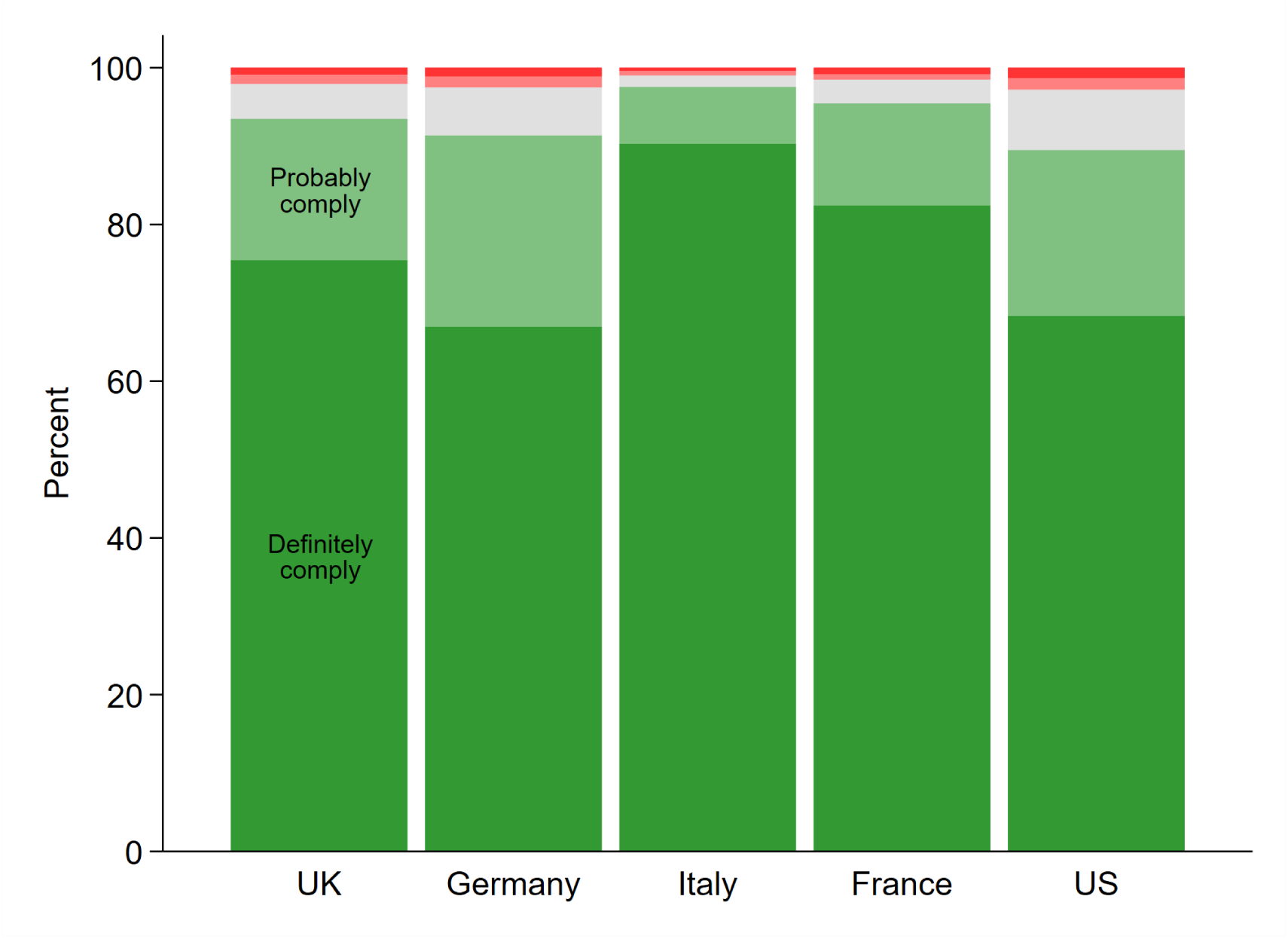

#### C.9 Effect of comprehension

In the German replication survey, conducted via Forsa (see Section B.3), we collected all answers also for those respondents who failed to answer the comprehension questions correctly. For this survey, we can thus compare the willingness to install between respondents who did (N=1048) and who did not (N=149) answer all comprehension questions correctly. We find that 69.0% of the respondents who answered the comprehension questions correctly would definitely or probably install the app. In contrast, only 56.4% of the respondents who answered wrongly would do so. This might be because the respondents who failed the comprehension questions answered randomly. Or it could be that these respondents are actually less willing to install.

The latter interpretation is consistent with previous literature[23] that showed a higher willingness to share data and information among the more technically informed users. A lack of understanding leading to a lower willingness to install would also be in line with the results of surveys that were conducted in Germany shortly after our survey. Some of these surveys did not explain the app in as much detail as our survey and these surveys often find lower willingness to install. For example, infratest Dimap asked respondents between 30 and 31 March 2020 about their willingness to install a contact-tracing app, and found that only 47% of respondents were willing to install the app.^9^ The comparison between our surveys and these shorter surveys suggests that a detailed explanation of how the app works and how it could mitigate or stop the epidemic increases the willingness to install.

### D Survey questionnaire

In the following, the UK version of the survey is presented with comments (in italics) explaining all the ways in which the Italian, French, German or US surveys differed from it. Overall, we tried to keep the surveys as similar as possible and only changed the phrasing if necessary (for example due to different political circumstances as some countries already were in lockdown at the time of the survey while others were not). In the US, we added a few more questions, which appear teal-colored in the overview below. Finally, horizontal lines indicate page breaks in the survey. The original survey texts for each country can be found here: UK, France, Germany, Italy and US.

#### D.1 Consent Form

In this study, we will ask you about an app that could help reduce the spread of the COVID-19 epidemic. You may ask any questions before deciding to take part by contacting the researchers (details below). The survey is about 10 minutes long. No background knowledge is required.

**Do I have to take part?** Please note that your participation is voluntary. If you do decide to take part, you may withdraw at any point during the survey for any reason before submitting your answers by closing the browser.

**How will my data be used?** Your answers will be completely anonymous. Your data will be stored in a password-protected file and may be used in academic publications. Research data will be stored for a minimum of three years after publication or public release.

**Who will have access to my data?** Lucid is the data controller with respect to your personal data and, as such, will determine how your personal data is used. Please see their privacy notice here: https://luc.id/privacy-policy/. Lucid will share only fully anonymised data with the University of Oxford, for the purposes of research. Responsible members of the University of Oxford and funders may be given access to data for monitoring and/or audit of the study to ensure we are complying with guidelines, or as otherwise required by law. This project has been reviewed by, and received ethics clearance through, the University of Oxford Central University Research Ethics Committee (reference number ECONCIA20-21-06).

**Who do I contact if I have a concern about the study or I wish to complain?** If you have a concern about any aspect of this study, please contact Johannes Abeler at and we will do our best to answer your query. We will acknowledge your concern within 10 working days and give you an indication of how it will be dealt with. If you remain unhappy or wish to make a formal complaint, please contact the Chair of the Research Ethics Committee at the University of Oxford who will seek to resolve the matter as soon as possible: Economics Departmental Research Ethics Committee at

**Please note that you may only participate in this survey if you are 18 years of age or over**.

If you have read the information above and agree to participate with the understanding that the data (including any personal data) you submit will be processed accordingly and that you need to be 18 years of age or over to participate, please confirm below.

○ I confirm
○ I do not confirm

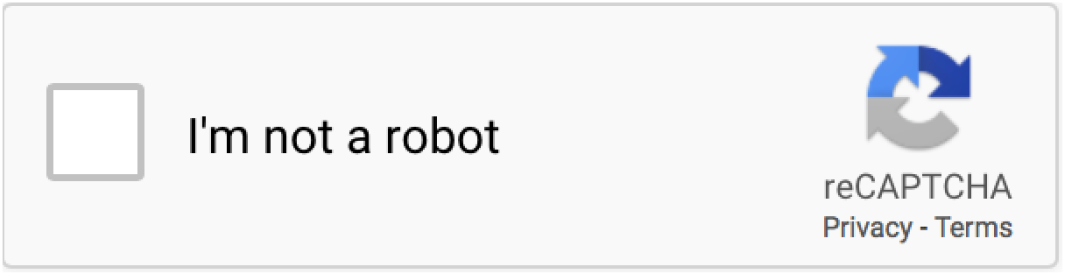

> In the US version, we asked the following filtering question on a separate screen:
>
> Is your area of residence currently under a stay-at-home order where you are no longer allowed to leave your home for non-essential reasons?
>
> ○ Yes
> ○ No
>
> Depending on the answer, participants continued with slightly different survey versions - one making reference to current restrictions and one referring to possible future restrictions. Below we will point out the few instances where this leads to minor differences in formulations.

#### D.2 App Description and Comprehension Questions

> The first screen was the same across all surveys, the only difference being that in the US survey we said “If enough people used the app, it could automatically alert you.”.

The current coronavirus epidemic (“COVID-19”) is all over the news.

People can get infected if they are in close contact with someone who has the virus. People do not notice when they get infected. They only notice when they start having a fever or a cough, perhaps a week later.

Imagine there was an app that you could install on your mobile phone. This app would automatically alert you if you had been in close contact for at least 15 minutes with someone who was infected with the coronavirus. Such an app does not exist yet in the UK. But we, researchers from the University of Oxford, are interested in understanding what you would think about such an app.

The next pages explain how such an app could work and will ask comprehension questions. You can only continue the survey if you answer all questions correctly.

> In the German, Italian and French versions, we said the app would only use Bluetooth and not location data. In every other country, we therefore listed “Activate Bluetooth” as the first (and correct) answer option in the comprehension question. Furthermore, we adapted the health services responsible for the app depending on the country, referring to the Center for Disease Control and Prevention (CDC) in the US, the Robert Koch-Institut (RKI) in Germany and more broadly to the “health services” in Italy and France.

The app would be developed by the NHS. You would need to install the app by simply clicking a link. Once installed, the app would register which other users are close to you. The app would do this by using Bluetooth and your location.

The app would NOT access your contacts, photos, or other data held on your phone. Only the NHS would have access to the data collected.

##### Comprehension check

What permission would you need to give the app?

- Permission to use my location data
- Permission to access my photos
- Permission to access my WhatsApp contacts

> Participants were only allowed to continue to the next screen if they selected option (1). If they chose one of the latter two options, the survey was terminated.
>
> In the US, Germany, Italy and France, we said the app would request people found to have been in contact with a confirmed case of COVID-19 to go into “quarantine at home” rather than “self-isolate” - terminology we stuck to throughout the respective surveys. We also explained the difference between the restrictions imposed on people in the existing lockdowns and the restrictions associated with being alerted by the app. In the UK, we did not consider this to be necessary as, at the time of the survey, the government had not issued a lockdown order yet. Finally, in the US survey, the “lockdown version” referenced a current stay-at-home-order while the “no lockdown version” mentioned a hypothetical one.

If the NHS diagnoses the coronavirus in somebody you have been in close contact with, the app would notify you automatically. The app would give you targeted advice on what to do. It will ask you to self-isolate at home for 14 days or until you have been tested for the virus.

This would be useful since people can infect others even before they have a fever or a cough. Self-isolating would thus protect your family, friends and colleagues from being infected by you. At the same time, only people who were in contact with an infected person would need to self-isolate.

If you had not been in close contact with a confirmed case, then the app would show you an “all clear” message.

##### Comprehension check

What would the app do if you were found to have been in contact with someone diagnosed with coronavirus?

- Ask me to self-isolate
- Give me an “all clear” message
- Tell me the name of the person who was diagnosed

> Participants were only allowed to continue to the next screen if they selected option (1). If they chose one of the latter two options, the survey was terminated.
>
> In the US, Italy, France and Germany, we stressed that, if enough people used it, the app could also shorten the duration of existing lockdowns (or school closures in the case of the “no-lockdown” US version).

If you are diagnosed with coronavirus, the app would notify all people you have been in close contact with, without identifying you to them, and advise them to self-isolate. This would increase the chance of finding all the people you might have infected and help make sure they can keep their loved ones safe as well. If enough people use the app, it will slow down the epidemic and might even stop it entirely.

##### Comprehension check

What would the app do if you were diagnosed with the coronavirus? o Give my name and address to all people I have been in close contact with

- Advise all people I have been in close contact with to self-isolate
- Shut down my phone

> Participants were only allowed to continue to the next screen if they selected option (2). If they choose the first or the last option, the survey was terminated.

#### D.3 Installation Questions

##### D.3.1 Voluntary Installation - General

> The general installation questions were the same in all countries. However, in the US, half of the participants were randomly assigned to see all the answer options in the survey in ascending rather than descending order i.e., the answer options were ordered from “Definitely won’t install” to “Definitely install” (with “Don’t know” remaining the last option). Whether or not a participant saw the ascending or descending order remained the same throughout the survey.

For the following questions, please imagine that an app like the one described before exists.

How likely would you be to install, or not install, the app on your phone?

- Definitely install
- Probably install
- May or may not install
- Probably won’t install
- Definitely won’t install
- Don’t know

> The ordering of the different reasons (both for and against installation) was randomized and people could choose multiple answers. In the non-UK versions, we gave one more reason for installing the app: “It would allow me to return more quickly to a normal life”. This is because in every country but the UK, the government had already issued a (partial) lockdown order.

What would be your main reasons for installing the app (you may click up to five)?

□ It would help me stay healthy
□ It would let me know my risk of being infected
□ Seeing the “all clear” message would give me peace of mind
□ It would protect my family and friends
□ It would help reduce the number of deaths among older people
□ A sense of responsibility to the wider community
□ It might stop the epidemic
□ Other (please indicate in the field below):

> In Italy, we gave “I worry the government would use this as an excuse for greater surveillance during the epidemic” (not just after) as an additional reason against, while in the US, we gave “I don’t believe other people will install it” as an additional reason against. Plus, in every non-UK country, we used “I don’t want to activate Bluetooth” rather than “I don’t want the [health services] to have access to my location data” as a reason against.

What would be your main reasons against installing the app (you may click up to five)?

□ The app would be too much hassle to install
□ I would not benefit from the app
□ I don’t want the NHS to have access to my location data
□ I won’t be infected anyway
□ I worry the government would use this as an excuse for greater surveillance after the epidemic
□ I don’t want to feel more anxious than I already feel
□ I worry that my phone will be more likely to get hacked
□ Other (please indicate in the field below):

How likely would you be to comply with the recommendation of the app to self-isolate at home for 14 days if you had been in close contact with an infected person?

○ Definitely comply
○ Probably comply
○ May or may not comply
○ Probably won’t comply
○ Definitely won’t comply
○ Don’t know

> Only participants who did NOT select “Definitely comply” in response to the previous question saw this screen.

Would you be more, or less, likely to comply with the advice to self-isolate for 14 days if the NHS committed to test you for the virus within 2 days from the start of your self-isolation? If you tested negative, you could stop self-isolating immediately.

- More likely
- Equally likely
- Less likely

> In the US version, participants were randomly assigned to see either a question about Mark Zuckerberg endorsing the app in a letter to all users or Facebook promoting the app on their website.

Would you be more, or less likely, to install such an app, developed by the CDC, if a private company like Facebook officially endorsed it, for example in a letter from their CEO Mark Zuckerberg?

- More likely
- Equally likely
- Less likely
- I don’t have a Facebook account

Would you be more, or less likely, to install such an app, developed by the CDC, if a private company like Facebook promoted it on their website?

- More likely
- Equally likely
- Less likely
- I don’t have a Facebook account

##### D.3.2 Voluntary Installation - Specific

Participants were only shown this entire section if they did not select “Definitely install” in response to the question “How likely would you be to install, or not install, the app on your phone?” at the beginning of the main questionnaire.

Many people in the UK worry about the effect of the virus on their community and on their family and friends.

Suppose someone in your community had been infected with the virus. How likely would you then be to install, or not install, the app on your phone?

- Definitely install
- Probably install
- May or may not install
- Probably won’t install
- Definitely won’t install
- Don’t know

> Only Participants who did not select “Definitely install” in response to the previous question were shown this screen.

Now suppose someone you personally know had been infected with the virus. How likely would you then be to install, or not install, the app on your phone?

- Definitely install
- Probably install
- May or may not install
- Probably won’t install
- Definitely won’t install
- Don’t know

> In Italy, France and Germany as well as areas under a stay-at-home order in the US, we dropped the “Imagine the government would introduce..” intro and instead only said: “Imagine the government decided to lift the restrictions of the current lockdown for those people for whom the app showed an all clear message. This means they would be able to leave their homes even without an essential reason.”

Imagine the government decided to introduce a full lockdown as in Italy to limit the spread of the coronavirus. This would mean that only essential stores, like supermarkets, would remain open, and you would only be allowed to leave your house in exceptional circumstances. Imagine that this restriction would be lifted for those people for whom the app showed an “all clear” message.

In this situation, how likely would you be to install, or not install, the app on your phone?

- Definitely install
- Probably install
- May or may not install
- Probably won’t install
- Definitely won’t install
- Don’t know

#### D.4 Automatic Installation

> In every other country, we stressed that people would be able to **immediately** uninstall the app - both in the intro and the first question.

Now, imagine the government asked the mobile phone providers (Vodafone, EE, etc.) to automatically install the app on all phones. This would maximise the chance of stopping the epidemic. You would be able to uninstall the app yourself and the app would automatically uninstall once the NHS has declared the epidemic to be over.

How likely would you be to keep, or uninstall, the app when the app is installed automatically?

- Definitely keep
- Probably keep
- May or may not keep
- Probably uninstall
- Definitely uninstall
- Don’t know

“The government should ask mobile phone providers to automatically install the app on all phones.” To what extent do you agree, or not, with the above statement?

- Fully agree
- Somewhat agree
- Neither agree nor disagree
- Somewhat disagree
- Fully disagree

> Only participants who did not select “Fully agree” in response to the previous question saw this screen.

“The government should ask mobile phone providers to automatically install the app on all phones.”

Now suppose someone in your community had been infected with the virus. To what extent do you agree, or not, with the above statement?

- Fully agree
- Somewhat agree
- Neither agree nor disagree
- Somewhat disagree
- Fully disagree

> Only participants who did not select “Fully agree” in response to either of the two previous questions saw this screen.

“The government should ask mobile phone providers to automatically install the app on all phones.”

Now suppose someone you personally know had been infected with the virus. To what extent do you agree, or not, with the above statement?

- Fully agree
- Somewhat agree
- Neither agree nor disagree
- Somewhat disagree
- Fully disagree

At the end of the epidemic, a decision would need to be made about what to do with the data collected. Which of the following policies would you prefer?

- All data will be automatically deleted at the end of the epidemic and not used for any other purpose
- All data will be de-identified and made available to university researchers to prepare for future epidemics
- Other (please indicate in the field below):

#### D.5 Demographics

How old are you?

- 18-30
- 31-40
- 41-50
- 51-60
- 61-70
- 71-80
- Older than 80

What is your gender?

- Female
- Male
- Other
- Prefer not to say

> In the US version, we asked an additional question about ethnicity.

What is your ethnicity (you may choose more than one)?

□ White
□ Asian
□ Black/African-American
□ Hispanic/Latino Origin
□ American Indian/Alaska Native
□ Other ethnicity or origin (please specify below):
□ Prefer no to say

> In all countries except the UK, we asked about state or region of residence, while in the UK, we used a higher level of aggregation.

Where in the UK do you currently reside?

- London
- England not including London
- Wales o Scotland
- Northern Ireland

> In the US version, we asked an additional question about the environment participants resided in.

How would you best describe the area you currently reside in?

- Urban
- Suburban
- Rural

> In all other countries, we replaced “socially” with “while getting groceries, working out, etc.”.

How often are you currently in close contact with people outside of your household, for example, at work or socially?

- Not more than once per week
- A few times per week
- A few times per day
- Many times per day

Do you have any of the following health problems: diabetes, high blood pressure, heart or breathing problems?

- Yes
- No

> In the US version, we asked an additional question about health insurance.

Do you currently have health insurance?

- Yes, I have private health insurance (e.g. by the employer)
- Yes, I have public health insurance (e.g. Medicare)
- No, I don’t have health insurance

How often do you have your mobile phone with you when you leave the house?

- Always
- Most of the time
- Sometimes
- Rarely

> In Italy, France, Germany and for areas under a lockdown in the US, we dropped the “Imagine…” intro and instead asked “How much work/study are you able to do from home, e.g., over the phone or the internet during this lockdown?”.

Imagine you would have to stay home for a week instead of going to work or to study, how much work/study would you be able to do from home, e.g., over the phone or the internet?

- None of my normal work
- About a quarter of my normal work
- About half of my normal work
- About three quarters of my normal work
- All of my normal work
- I do not work or study

Would you receive sick pay or continue to receive your income if you stayed and worked from home?

- Yes
- No
- Don’t know

> In Italy, France and Germany, we asked “Which of the following political parties do you feel closest to?”. In the US, we asked which party respondents voted for in the last presidential election and displayed the question on a separate screen, while in the other versions it was on the same screen as the “trust in government” question.

Do you think of yourself as a supporter of any one of the following political parties?

- Conservatives
- Liberal Democrats
- Labour
- Brexit Party
- SNP
- Plaid Cymru
- Other or I don’t want to say

> In the US version, we asked participants about their main source of information.

Where do you get most of your news from?

- Newspaper
- Television
- Radio
- Family & Friends
- Social Media

> US Participants only saw this screen if they selected “Television” as their main source of information in the question about news.

Which television channel do you watch most frequently for the news?

- CNN
- MSNBC
- Fox News
- ABC
- CBS
- Other (Please specify below):

> US Participants only saw this screen if they selected “Newspaper” as their main source of information in the question about news.

Which newspaper do you read most frequently (either in print or online)?

- The New York Times
- Washington Post
- New York Post
- USA Today
- he Wall Street Journal
- Los Angeles Times
- Other (Please specify below):

> US Participants only saw this screen if they selected “Social media” as their main source of information in the question about news.

Which social media platform do you get most of your news from?

- Facebook
- Instagram
- Twitter
- LinkedIn
- WhatsApp
- Other (Please specify below):

To what extent do you agree with the following statement: “I generally trust the government to do what is right.”?

- Fully agree
- Somewhat agree
- Neither agree nor disagree
- Somewhat disagree
- Fully disagree

#### D.6 Political Assessment

To what extent do you agree, or not, with the following statement: “My opinion about the British government would improve if they introduced such an app and allowed me to decide myself whether to install it or not.”?

- Fully agree
- Somewhat agree
- Neither agree nor disagree
- Somewhat disagree
- Fully disagree

To what extent do you agree, or not, with the following statement: “My opinion about the British government would improve if they asked mobile phone providers to automatically install such an app on all phones to maximise the chance of stopping the epidemic.”?

- Fully agree
- Somewhat agree
- Neither agree nor disagree
- Somewhat disagree
- Fully disagree

> In the US version, we asked an additional, direct question about which installation regime respondents would prefer.

Imagine that the federal government introduced such an app - which of the two installation regimes described above would you prefer?

- Voluntary installation
- Automatic installation (with an option to uninstall)

#### D.7 Debrief

> In each country, we referred participants to the coronavirus information website of the relevant government agency.

Thank you very much!

If you have any questions about the app or any feedback, please let us know by writing them into the field below. You can also email the researchers at.

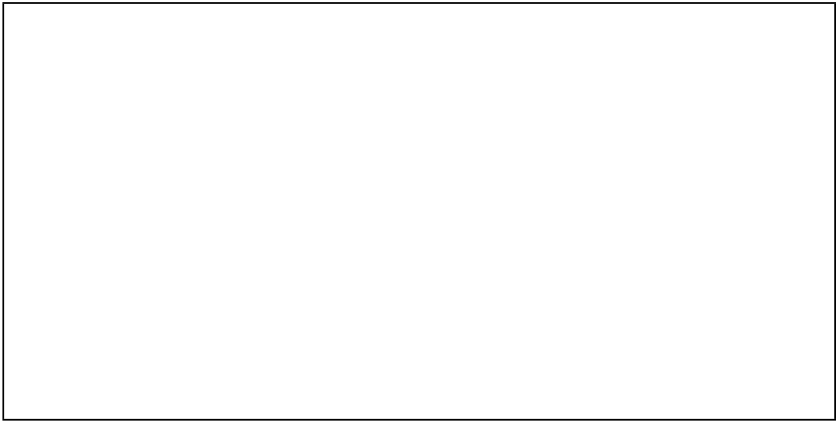

If you want to know more about the coronavirus and how to protect you and your family, please click this link to the NHS coronavirus website: https://www.nhs.uk/coronavirus

Please click the button below to finish the survey.

## Footnotes

1 Respondents could also select the option *Don’t know*, which in the analysis we merge with the mid-point of the scale, *May or may not install*. The options on the scale were always presented in descending order, except in the US where we randomized whether respondents saw the scale in descending or ascending order to test for possible order effects. We found no evidence of this. See Section D in the Multimedia Appendix for more information.

2 For example, in the UK and areas in the US that were not under lockdown, we asked about willingness to install if the (local) government firstly introduced a lockdown and then tied a relaxation of restrictions to an “all clear” message on the app. In contrast, in Germany, Italy, France and areas in the US under a stay-at-home order, we simply asked about willingness to install if current restrictions were lifted in response to an “all clear” message.

3 A few respondents still noted that they did not have a smartphone in the comments section (as a reason for not installing the app) - we excluded those responses from our final sample.

4 For the few people who took the survey more than once, we kept their first submission.

5 Because there were so few people who either wanted to keep their gender private or identified as non-binary, it did not seem useful to have an extra category when analyzing the results by gender.

6 To recruit participants, Forsa randomly generates phone numbers, calls the respective households and invites them to participate in the survey. Due to the direct contact over the phone, the sample should contain neither bots, nor respondents with a dubious identity (due to fake identities or multiple sign-ups).

7 The validity of using these inferred preferences is considered in Multimedia Appendix subsection C.6.1.

8 As this survey asks hypothetical questions, we should caveat that *actual* compliance might be considerably lower.

9 The exact question (translated from German) was: \Assume that there is an app for your mobile phone that would allow you to track your COVID-19 symptoms as well as document your geolocation data. The app could thus show all users whether they had been close to a person infected with the coronavirus. Would you use such an app or not?”

